# Holistic AI-Driven Quantification, Staging and Prognosis of COVID-19 Pneumonia

**DOI:** 10.1101/2020.04.17.20069187

**Authors:** Guillaume Chassagnon, Nikos Paragios

## Abstract

Improving screening, discovering therapies, developing a vaccine and performing staging and prognosis are decisive steps in addressing the COVID-19 pandemic. Staging and prognosis are especially crucial for organizational anticipation (intensive-care bed availability, patient management planning) and accelerating drug development; through rapid, reproducible and quantified response-to-treatment assessment. In this letter, we report on an artificial intelligence solution for performing automatic staging and prognosis based on imaging, clinical, comorbidities and biological data. This approach relies on automatic computed tomography (CT)-based disease quantification using deep learning, robust data-driven identification of physiologically-inspired COVID-19 holistic patient profiling, and strong, reproducible staging/outcome prediction with good generalization properties using an ensemble of consensus methods. Highly promising results on multiple independent external evaluation cohorts along with comparisons with expert human readers demonstrate the potentials of our approach. The developed solution offers perspectives for optimal patient management, given the shortage of intensive care beds and ventilators^1, 2^, along with means to assess patient response to treatment.

## Main

COVID-19 emerged in December 2019 in Wuhan, China^3^ – its worldwide spread lead the World Health Organization to declare the COVID-19 outbreak as a pandemic on March 11th, 2020. The disease, caused by the SARS-Cov-2 virus has respiratory failure due to severe viral pneumonia^4^ as the leading cause of death. Key to addressing the COVID-19 pneumonia pandemic is the ability to perform - both continuously and upon hospital admission - quantification, staging and short/long-term prognosis. Identifying patients who are most likely to deteriorate and require intubation could accord efficient management of patient population and hospital resources, through anticipation regarding transfer to other hospitals, and selection regarding patients for new drugs, thus reducing the need for intubation. To the best of our knowledge, there is currently no validated model able to predict short- or long-term outcome, barring poor-outcome risk factors such as age, sex, disease extent, several biological biomarkers and comorbidities^4–10^.

In this study, our clinical objective is three-fold:

- fully automatic disease quantification from CT scans, facilitating severity estimation towards optimal patient care and faster drug trials outcomes assessment.
- COVID19-specific holistic, highly compact multi-omics explainable patient signature integrating imaging/clinical/biological data and associated comorbidities for automatic patient staging.
- short and long-term prognosis for clinical resources optimization offering alternative/complementary means to facilitate triage.

Artificial Intelligence (AI) aims either to reproduce human behavior with respect to a specific task (a given set of observations and the corresponding experts’ assessments) or to find and better understand correlations between input signals and outcomes (invisible to the human eye). AI has gained tremendous attention in recent years in medical health, providing valuable tools toward diagnosis, treatment and personalization^11–13^. AI, more recently, contributed to understanding new diseases such as COVID-19^14,15^. In particular, studies already report the use of artificial intelligence in distinguishing COVID-19 patients from community-acquired pneumonia on CT^16^, in computing the protein structures associated with COVID-19, or even in discovering genomic signatures for the rapid classification of COVID-19 patients^17^. While CT has rapidly assumed a major role for COVID-19 diagnosis^18^, it is also used for staging and severity assessment along with known biological and clinical biomarkers such as D-dimer levels and lymphocyte count or obesity, diabetes and hypertension indicators^4,7,9,19^.

In this study we investigated an automatic method (Figure 1) for COVID-19 pneumonia quantification, staging and prognosis that integrates known biological, clinical and self-discovered interpretable CT-imaging biomarkers. Our rationale was that imaging and biological/clinical information are complementary - their holistic integration bearing novel means of understanding the disease. Our approach relied on (i) a disease quantification solution that exploited an ensemble of 2D & 3D convolutional neural networks, (ii) a confident biomarker discovery approach seeking to determine the shared compact space of imaging, biological and clinical features that are the most COVID19 informative, (iii) an ensemble consensus-driven robust, reproducible, explainable AI method to reliably perform staging and prognosis.

**Figure 1.**
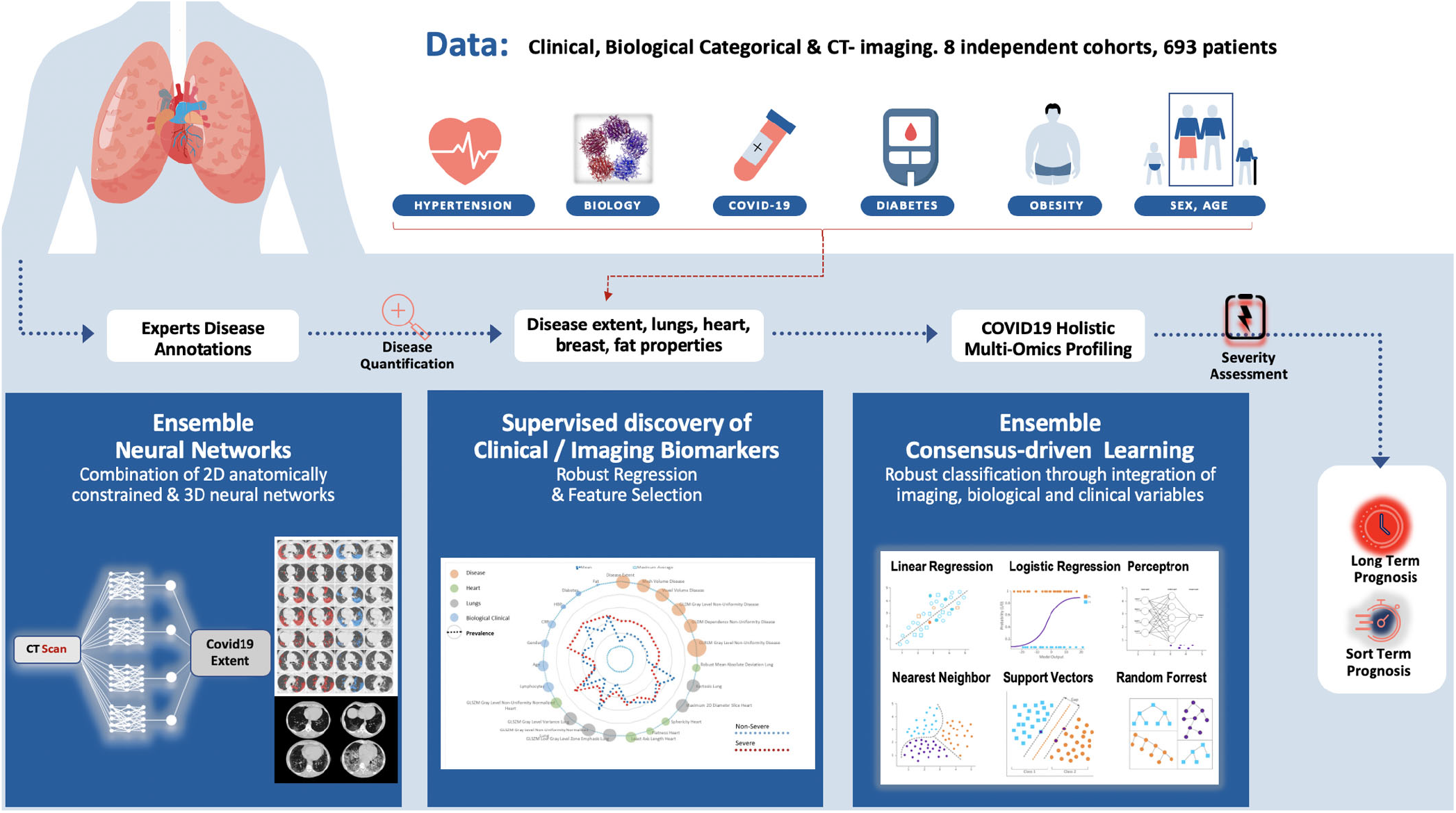
Overview of the method for CT-based quantification, staging and prognosis of COVID-19. (i) Two independent cohorts with quantification based on ensemble 2D & 3D consensus neural networks reaching expert-level annotations on massive evaluation, (ii) Consensus-driven COVID19 bio(imaging)-holistic profiling and staging & (iii) Consensus of linear & non-linear classification methods for short and long term prognosis.

### Part I: Disease Quantification

We report on a deep learning-based segmentation tool to quantify the COVID-19 disease and lung volume, using an ensemble network approach combining a 2D (AtlasNet framework^20^) and a 3D (3D-UNet^21^) architecture. We investigated a combination of 2D slice-based^20,22^ and 3D patch-based ensemble architectures^21^. The development of the deep learning-based segmentation solution was done on the basis of a multi-centric cohort of unenhanced chest CT scans performed at initial evaluation of COVID-19 patients with positive Reverse transcription polymerase chain reaction (RT-PCR). The multicentric dataset was acquired at 6 hospitals, equipped with 4 different CT models from 3 different manufacturers, with different acquisition protocols and radiation doses (Table 1). Fifty CT exams from three centers were used for training and 130 CT exams from three different centers were used for test (Table 2). Disease and lung were delineated on all 23, 423 slices used of the training dataset, and on only 20 slices – equispaced, covering the entire lung region per exam - but by 2 independent annotators in the test dataset (2, 600 images). The overall annotation effort took approximately 800 hours and involved 15 radiologists with 1 to 7 years of experience in chest imaging.

**Table 1.** Acquisition and reconstruction parameters. *Note*.*— For quantitative variables, data are mean* ± *standard deviation, and numbers in brackets are the range. CT = Computed Tomography* ; *CTDIvol = volume Computed Tomography Dose Index* ; *DLP = Dose Length Product * significant difference with p* < 0.001

|  | Center A | Center B | Center C | Center D | Center E | Center F | Center G | Center H |
| --- | --- | --- | --- | --- | --- | --- | --- | --- |
| CT equipment | Somatom AS+ | Resolution HD | Aquilion Prime | Somatom Edge | Revolution HD | Aquilion Prime | Revolution HD | Somatom AS+ |
| Kilovoltage | 100-120 | 120 | 100-120 | 100-120 | 120-140 | 100-120 | 120 | 100-120 |
| DLP (mGy.cm) | 109 ± 42<br>[44-256] | 306 ± 104<br>[123-648] | 102 ± 30<br>[43-189] | 131 ± 44<br>[55-499] | 177 ± 48<br>[43-276] | 115 ± 26<br>[75 - 186] | 285 ± 108<br>[70 - 679] | 332 ± 156<br>[179 - 755] |
| CTDIvol (mGy) | 3.2 ± 1.5<br>[1.2-11.9] | 8.7 ± 2.8<br>[3.9-18.5] | 2.7 ± 0.9<br>[1.0-5.3] | 3.2 ± 0.9<br>[1.4-9.5] | 5.5 ± 1.8<br>[1.2-12.3] | 2.5 ± 0.6<br>[1.7-4.3] | 7.9 ± 2.9<br>[1.7-18.0] | 8.5 ± 4.0<br>[4.4-19.8] |
| Slice thickness | 1mm | 0.625mm | 1mm | 0.625mm | 1mm | 1mm | 0.625mm | 1mm |
| Convolution Kernel | i70 | Lung | FC51-FC52 | i50 | Lung | FC51-FC52 | Lung | i70 |
| Iterative reconstructions | SAFIRE 3 | ASIR-v 80% | IDR 3D0.67 | SAFIRE 4 | ASIR-v 60% | IDR 3D | ASIR-v 60% | SAFIRE 3 |

**Table 2.** Patient characteristics in the datasets used for developing the segmentation tool. *Note. For quantitative variables, data are mean* ± *standard deviation, and numbers in brackets are the range. CT = Computed Tomography; CTDIvol = volume Computed Tomography Dose Index; DLP = Dose Length Product*

|  | Training/Validation Dataset<br>(Centers A+B+C; N=50) | Test Dataset<br>(Centers D+E+F; n=130) | p value |
| --- | --- | --- | --- |
| Age (y) | 57 ± 17 [26-97] | 59 ± 16 [17-95] | 0.363 |
| No. of Men | 31(62) | 87(67) | 0.534 |
| Disease extent* |  |  |  |
| Manual | 18.1 ± 14.9 [0.3-68.5] | 19.5 ± 16.5 [1.1-75.7] | 0.574 |
| Automated | - | 19.9% ± 17.7 [0.5-73.2] | - |
| DLP (mGy.cm) | 180 ± 124 [43-527] | 139 ± 49.0 [43-276] | 0.026 |
| CTDIvol (mGy) | 4.9 ± 3.4 [1.0-13.0] | 4.0 ± 1.9 [1.2-12.3] | 0.064 |

The consensus between manual (2 annotators) and AI segmentation was measured using the Dice similarity score (DSC)^23^ and the Hausdorff distance (HD). The CovidENet performed equally well to trained radiologists in terms of DSCs and better in terms HD (Figure 2, 5). The mean/median DSCs between the two expert annotations on the test dataset were 0.70*/*0.72 for disease segmentation while DSCs between CovidENet and the manual segmentations were 0.69*/*0.71 and 0.70*/*0.73. In terms of HDs, the average expert distance was 9.16mm while it was 8.96mm between CovidENet and the experts. When looking at disease extent (percentage of lung affected by the disease) no significant difference was observed between the AI and the manual segmentations’ average (19.9% ± 17.7[0.5 − 73.2] vs 19.5% ± 16.5[1.1 − 75.7]; p= 0.352).

**Figure 2.**
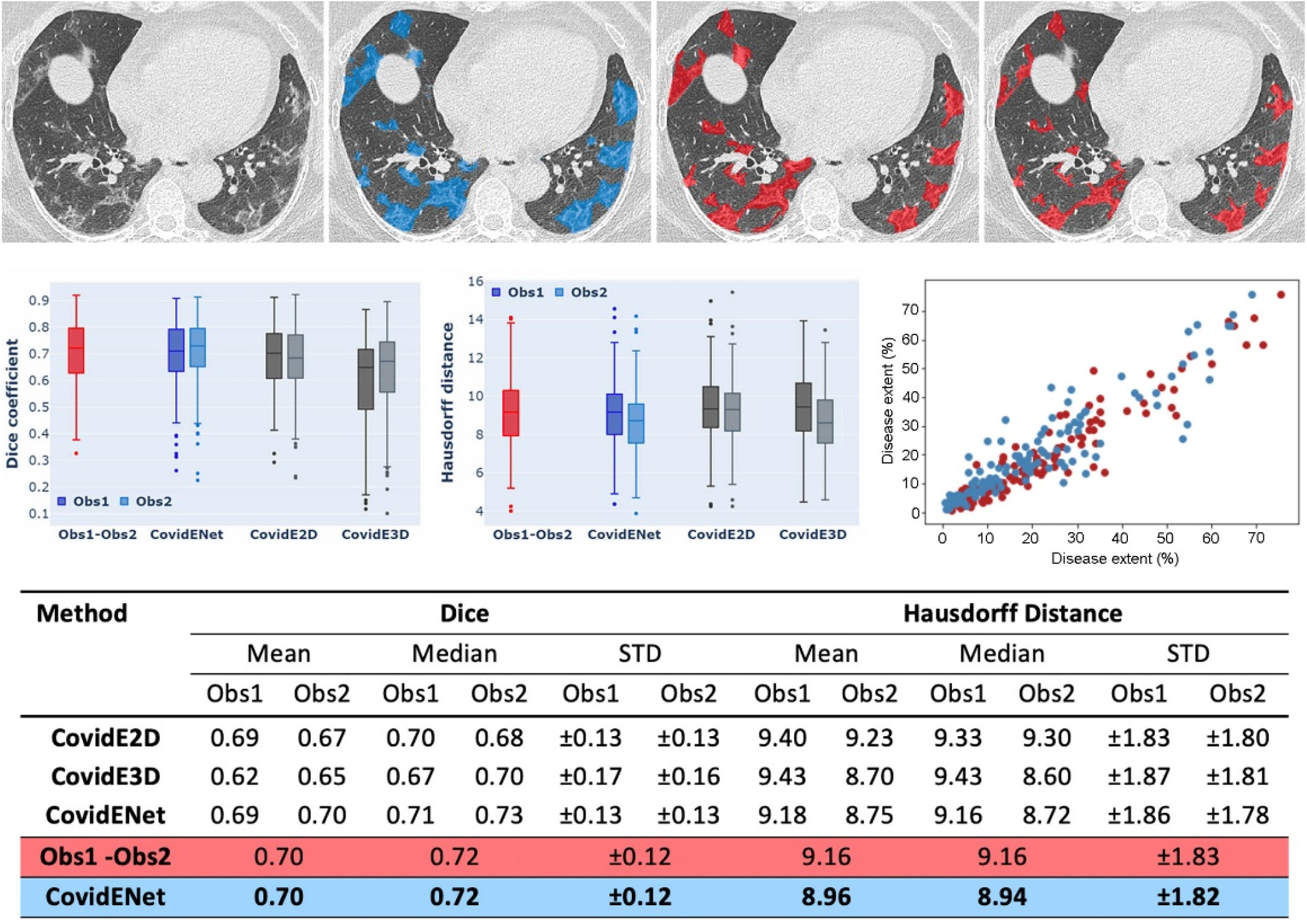
Comparison between automated and manual segmentation. Delineation of the diseased areas on chest CT in a COVID-19 patient: First Row: input, AI-segmentation, expert I-segmentation, expert II-segmentation. Second Row: Box-Plot Comparisons in terms of Dice similarity and Haussdorf between AI-solution, expert I & expert II, & Plot of correlation between disease extent automatically measured and the average disease extent measured from the 2 manual segmentation. Disease extent is expressed as the percentage of lung affected by the disease. Third row: statistical measures on comparisons between AI, expert I, and expert II segmentation.

### Part II: COVID-19 Holistic Multi-Omics Profiling & Staging

Multidisciplinary medical expertise endowed with clinical, biological, pathological, imaging patient’s data are essential to optimize and standardize COVID-19 pneumonia patient care. In a pandemic, such objectives are elusive due to (i) limited understanding of the disease, (ii) fast progression in terms of symptoms, and (iii) lack of qualified practitioners, primarily concentrated at urban centers. AI can address these limitations in the context of a pandemic. Automatic analysis of clinical, biological and imaging data through an evidence-driven approach can lead to the development of a compact holistic signature to assess the severity of the disease and provide an estimate of disease-free survival at the hospital admission stage. Our study reviewed outcomes in patient charts within the 4 (short-term) and 31+ days (long-term) at admission, dividing the patients in 2 groups: patients who died, or required mechanical ventilation either at initial or a subsequent admission as severe cases (S), and patients who were non-severe cases (NS). Two disjoint independent sets were built for training (536 patients from 5 centers) and testing (157 patients from the remaining 3 centers) (Table 3). COVID-19 multidisciplinary, multi-omics patient profiling was achieved through an evidence-driven mining approach.

**Table 3.**
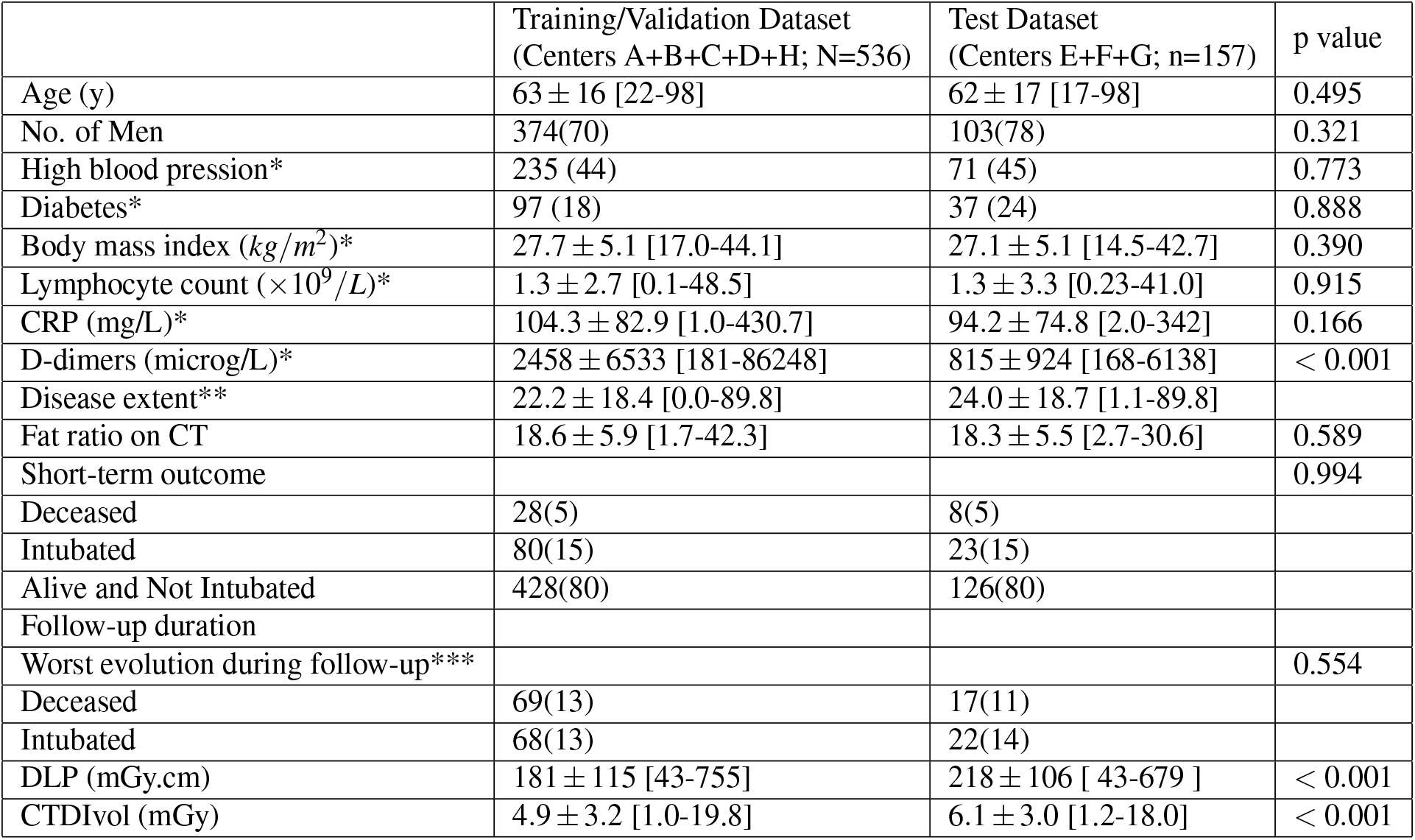
Patient characteristics of the training and testing datasets used for the creation of the Holistic Multi-Omics COVID19 signature and the development of short, long-term prognosis tools. *Note*.*— For quantitative variables, data are mean* ± *standard deviation, and numbers in brackets are the range. For qualitative variables, data are numbers of patients, and numbers in parentheses are percentages. CT = computed tomography, CTDIvol = volume Computed Tomography Dose Index; DLP = Dose Length Product * Available clinical data: n* = 692 *for diabetes and high blood pressure*(*leading to* 0.19*% of missing data on the training set*), *n* = 674 *for lymphocyte count* (*leading to* 2.05*% and* 5.10*% of missing data on the training and test sets respectively*), *n* = 654 *for CRP* (*leading to* 4.66*% and* 8.92*% of missing data on the training and test sets respectively*), *n* = 362 *for Body Mass Index, and n* = 339 *for D-dimers. **Percentage of lung volume on the whole CT, ***data available for 688 patients*

|  | Training/Validation Dataset<br>(Centers A+B+C+D+H; N=536) | Test Dataset<br>(Centers E+F+G; n=157) | p value |
| --- | --- | --- | --- |
| Age (y) | 63 ± 16 [22-98] | 62 ± 17 [17-98] | 0.495 |
| No. of Men | 374(70) | 103(78) | 0.321 |
| High blood pressure* | 235 (44) | 71 (45) | 0.773 |
| Diabetes* | 97 (18) | 37 (24) | 0.888 |
| Body mass index ($kg/m^2$)* | 27.7 ± 5.1 [17.0-44.1] | 27.1 ± 5.1 [14.5-42.7] | 0.390 |
| Lymphocyte count ($\times 10^9/L$)* | 1.3 ± 2.7 [0.1-48.5] | 1.3 ± 3.3 [0.23-41.0] | 0.915 |
| CRP (mg/L)* | 104.3 ± 82.9 [1.0-430.7] | 94.2 ± 74.8 [2.0-342] | 0.166 |
| D-dimers (microg/L)* | 2458 ± 6533 [181-86248] | 815 ± 924 [168-6138] | < 0.001 |
| Disease extent** | 22.2 ± 18.4 [0.0-89.8] | 24.0 ± 18.7 [1.1-89.8] |  |
| Fat ratio on CT | 18.6 ± 5.9 [1.7-42.3] | 18.3 ± 5.5 [2.7-30.6] | 0.589 |
| Short-term outcome |  |  | 0.994 |
| Deceased | 28(5) | 8(5) |  |
| Intubated | 80(15) | 23(15) |  |
| Alive and Not Intubated | 428(80) | 126(80) |  |
| Follow-up duration |  |  |  |
| Worst evolution during follow-up*** |  |  | 0.554 |
| Deceased | 69(13) | 17(11) |  |
| Intubated | 68(13) | 22(14) |  |
| DLP (mGy.cm) | 181 ± 115 [43-755] | 218 ± 106 [43-679] | < 0.001 |
| CTDIvol (mGy) | 4.9 ± 3.2 [1.0-19.8] | 6.1 ± 3.0 [1.2-18.0] | < 0.001 |

Information relevant to the disease volume and nature (CT-imaging), the non-COVID 19 affected lungs (CT-imaging), the heart condition / volume of the heart (CT-imaging), obesity (CT-imaging), biological data (lymphocytes, C-reactive protein (CRP) levels) and known demographic (age & sex)/confounding (diabetes, hypertension) factors were aggregated. Least absolute shrinkage and selection operator, with different classification methods, along with statistical significance variable selection methods led to the creation of low-dimensional holistic clinical, biological, CT-imaging COVID-19 patient multi-omics signature. This holistic COVID-19 pneumonia signature is presented in (Table 4) along with the prevalence of selection and the correlations with outcome. The average signature for the severe and non-severe case in the test set are presented in Figure 3. Consensus ensemble learning through majority voting was used to determine the subset of AI methods that have robust, reproducible performance with good generalization properties. Human “reader+” was used as a reference through consensus among three chest radiologists (resident, 7+ years of experience, 20+ years of experience in thoracic imaging). The AI solution aiming to separate patients with severe and non-severe short-term outcomes had a balanced accuracy of 70% (vs 67% for human readers consensus), a weighted precision of 81% (vs 78%), a weighted sensitivity of 64% (vs 70%) and specificity of 77% (vs 64%) (Figure 3, Table 5) and outperformed the consensus of human readers (Figure 3, Table 6). The AI solution successfully predicted 81% of the severe/critical cases opposed to only 61% for the consensus reader. Severe cases as depicted in Figure 3 referred to diabetic men, with higher level of volume/heterogeneity of disease and C-reactive protein levels.

**Table 4.** Supervised biomarker discovery for the creation of the Holistic Multi-Omics COVID19 signature. Each of the selected features together with their prevelances per feature selection method as well as on the final concensus is presented. *Note*.*— GLSZM =Gray Level Size Zone Matrix, GLRLM = Gray Level Run Length Matrix, GLDM = Gray Level Dependence Matrix*

| **Clinical** |  |  |  |  |  |  |  |  |  |
| --- | --- | --- | --- | --- | --- | --- | --- | --- | --- |
| Features | Lasso | Decision Tree | Linear SCV | XG Boost | AdaBoost | ch2 | ANOVA F-value | Mutual Information | Consensus |
| Lymphocytes | 0 | 0.25 | 0.66 | 1 | 0.57 | 0 | 0 | 0.02 | 0.31 |
| Age | 0.53 | 0.03 | 0.94 | 0.82 | 0.12 | 0 | 0 | 0.02 | 0.31 |
| Sex | 0.98 | 0 | 0.95 | 0 | 0 | 0.02 | 0 | 0.01 | 0.25 |
| CRP | 0 | 0.16 | 0.65 | 0.88 | 0.14 | 0 | 0 | 0 | 0.15 |
| HBP | 0.39 | 0 | 0.75 | 0.02 | 0 | 0 | 0 | 0 | 0.23 |
| Diabetes | 0.32 | 0 | 0.6 | 0.03 | 0 | 0.01 | 0 | 0 | 0.12 |
| **Indexes** |  |  |  |  |  |  |  |  |  |
| Features | Lasso | Decision Tree | Linear SVC | XG Boost | AdaBoost | chi2 | ANOVA F-value | Mutual Information | Consensus |
| Fat index | 0 | 0.04 | 0.93 | 0.42 | 0.06 | 0 | 0 | 0 | 0.18 |
| Disease Extent | 0.01 | 0.08 | 0.34 | 0.25 | 0.07 | 1 | 0.95 | 0.81 | 0.44 |
| **Heart** |  |  |  |  |  |  |  |  |  |
| Features | Lasso | Decision Tree | Linear SVC | XG Boost | AdaBoost | chi2 | ANOVA F-value | Mutual Information | Consensus |
| Maximum 2D Diameter Slice | 0.25 | 0.03 | 0.93 | 0.65 | 0.08 | 0 | 0 | 0 | 0.24 |
| Non-Uniformity on the GLSZM | 0.68 | 0 | 0.89 | 0.01 | 0 | 0 | 0 | 0 | 0.20 |
| Sphericity | 0 | 0.02 | 0.62 | 0.78 | 0.04 | 0 | 0 | 0 | 0.18 |
| Flatness | 0 | 0.13 | 0.88 | 0.34 | 0.1 | 0 | 0 | 0 | 0.18 |
| Least Axis Length | 0 | 0 | 0.81 | 0.29 | 0 | 0 | 0 | 0 | 0.14 |
| **Lung** |  |  |  |  |  |  |  |  |  |
| Features | Lasso | Decision Tree | Linear SVC | XG Boost | AdaBoost | chi2 | ANOVA F-value | Mutual Information | Consensus |
| Mean Absolute Deviation | 0.59 | 0 | 0.49 | 0.4 | 0 | 0.86 | 0.94 | 0.97 | 0.53 |
| Kurtosis | 0 | 0.72 | 0.92 | 0.98 | 0.73 | 0.07 | 0.01 | 0.64 | 0.51 |
| Zone Emphasis on the GLSZM | 0 | 0.02 | 0.92 | 0.5 | 0.01 | 1 | 0.82 | 0.45 | 0.47 |
| Non-Uniformity on the GLSZM | 1 | 0 | 0.98 | 0.49 | 0.02 | 0.01 | 0.92 | 0.26 | 0.46 |
| Variance on the GLSZM | 0 | 0 | 0.98 | 0.39 | 0.03 | 1 | 0.92 | 0.26 | 0.45 |
| **Disease** |  |  |  |  |  |  |  |  |  |
| Features | Lasso | Decision Tree | Linear SVC | XG Boost | AdaBoost | chi2 | ANOVA F-value | Mutual Information | Consensus |
| Non-Uniformity on the GLSZM | 0.95 | 0.18 | 0.88 | 0.91 | 0.23 | 1 | 1 | 0.93 | 0.76 |
| Non-Uniformity on the GLDM | 0.03 | 0.23 | 0.6 | 0.86 | 0.24 | 0.77 | 1 | 0.69 | 0.55 |
| Mesh Volume | 0.08 | 0.03 | 0.99 | 0.4 | 0.03 | 0.99 | 1 | 0.73 | 0.53 |
| Voxel Volume | 0.02 | 0 | 0.99 | 0.47 | 0.05 | 0.99 | 1 | 0.57 | 0.51 |
| Non-Uniformity on the GLRLM | 0 | 0.1 | 1 | 0.74 | 0.15 | 0.3 | 0.96 | 0.83 | 0.51 |

**Table 5.** Performances of each of the top-5 individual classifiers and of the ensemble classifier to differentiate between patient with Severe (S) and Non-Severe (NS) short-term outcome *Note*.*- L-SVM = Support Vector Machine with a linear kernel; RBF-SVM = Support Vector Machine with a Radial Basis Function kernel*

| Classifier | Balanced Accuracy |  | Weighted Precision |  | Weighted Sensitivity |  | Weighted Specificity |  |
| --- | --- | --- | --- | --- | --- | --- | --- | --- |
|  | Training | Test | Training | Test | Training | Test | Training | Test |
| L-SVM | 0.7 | 0.67 | 0.79 | 0.78 | 0.71 | 0.71 | 0.69 | 0.64 |
| RBF-SVM | 0.75 | 0.68 | 0.82 | 0.79 | 0.7 | 0.67 | 0.79 | 0.7 |
| Decision Tree | 0.71 | 0.67 | 0.82 | 0.82 | 0.61 | 0.53 | 0.81 | 0.81 |
| Random Forest | 0.72 | 0.68 | 0.81 | 0.79 | 0.69 | 0.69 | 0.75 | 0.68 |
| AdaBoost | 0.72 | 0.67 | 0.83 | 0.82 | 0.63 | 0.54 | 0.82 | 0.81 |
| Ensemble Classifier | 0.73 | 0.7 | 0.82 | 0.81 | 0.67 | 0.64 | 0.8 | 0.77 |

**Table 6.**
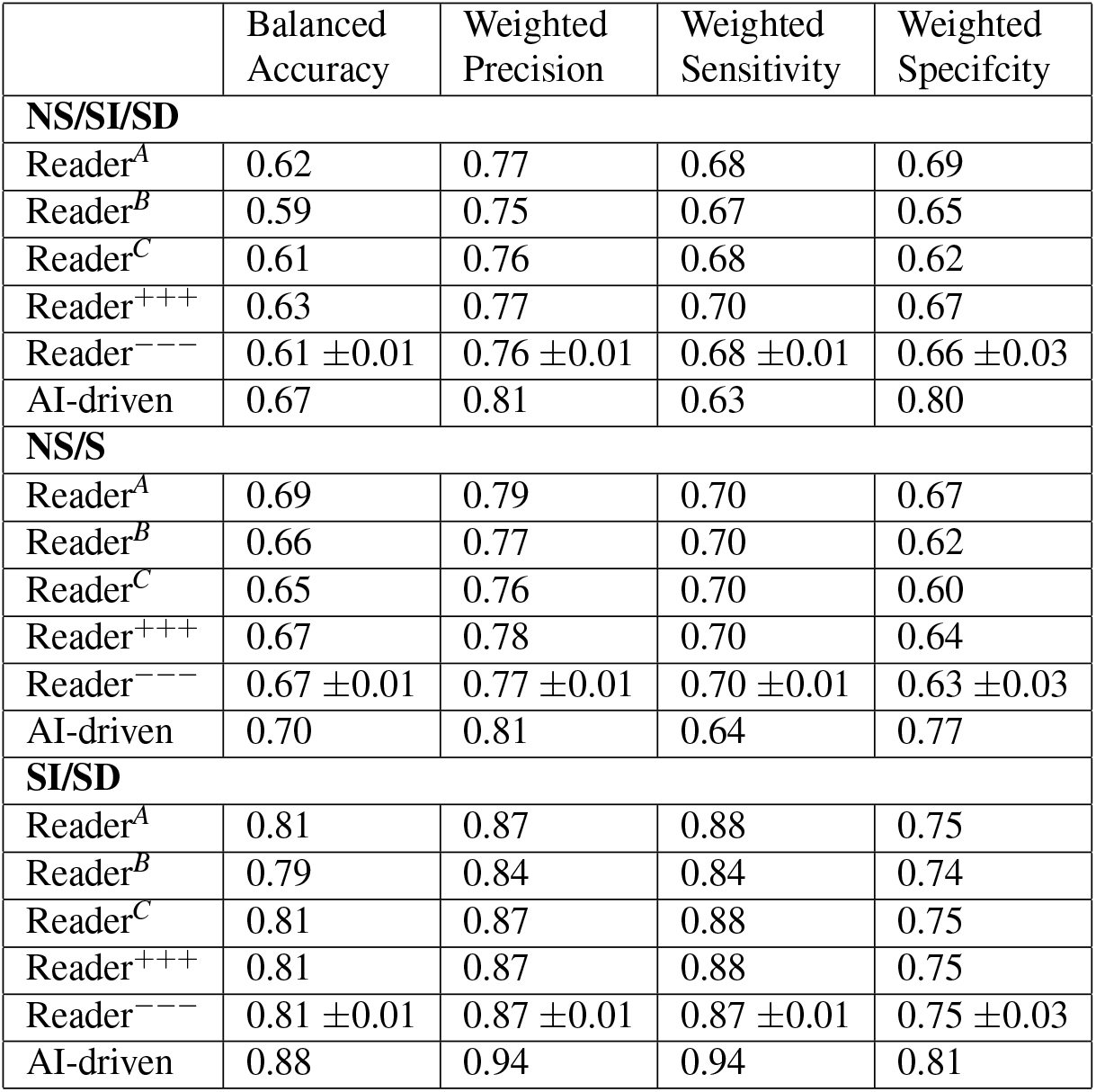
Prognosis of medical experts for the Non Severe (NS) versus Severe (S), Intubated (SI) versus Deceased (SD) and NS/SI/SD patients *Note*.*- Classification Performance Reader*^*A*^ (*Senior*), *Reader*^*B*^ (*Established*), *Reader*^*C*^ (*Resident*),*Reader*^+++^ (*Consensus among Human Readers*), *Reader*^−−−^ (*Average performance of Human Readers*)

**Figure 3.**
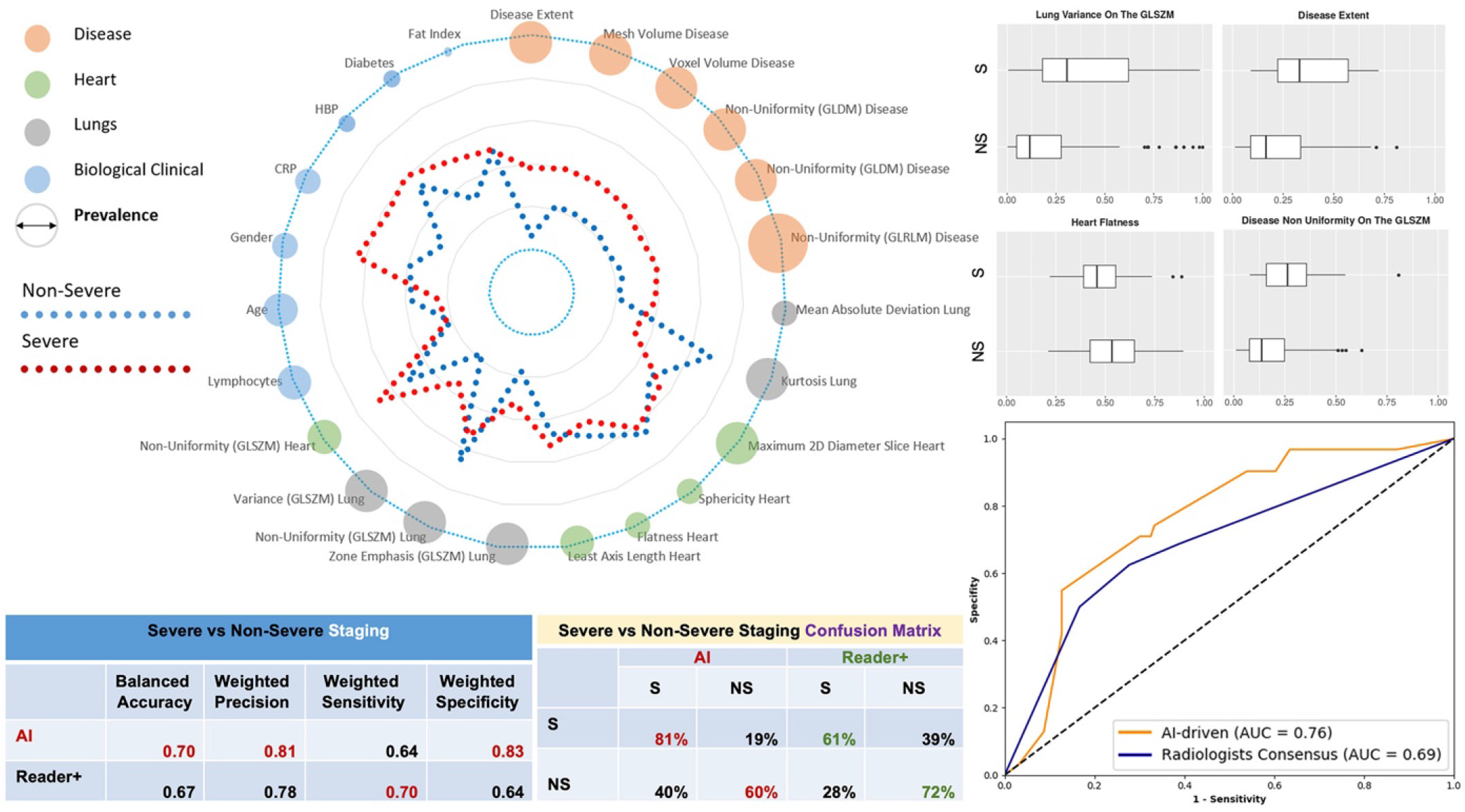
COVID-19 Holistic Multi-Omics Signature Staging: Average profiles (spider chart) with respect to the with respect to the severe versus non-severe separation are shown along with prevalence of biomarkers (diameter of the circle). Classification performance, confusion matrices and area under the curve with respect to the AI and the consensus of expert readers (reader+) are reported. Selective associations of features with outcome are shown (box plots).

### Part III: Short & Long-Term Prognosis

The COVID-19 pneumonia pandemic spiked hospitalizations, while exerting extreme pressure on intensive care units. In the absence of a cure, staging and prognosis is crucial for clinical decision-making for resource management and experimental outcome assessment, in a pandemic context. Our objective was to predict patient outcomes prior to mechanical ventilation support. Similar to staging, we reviewed outcomes of all patients and created three distinct subpopulations: those who had a short term negative (SD = short-term deceased) outcome (deceased within 4 days after admission), those who didn’t recover within 30 days of mechanical ventilation (LD= long-term deceased) and the ones who recovered within 30 days on mechanical ventilation (LR= long-term recovered). The data was separated in the same manner as for the staging task leading to a total of 139 patients with severe symptoms - 108 patients (5 centers) for training and 31 for testing (3 centers). Prognosis was addressed in a hierarchical manner, first targeting the separation between short-term negative and long-term outcomes, to differentiate positive and negative long-term outcomes. Consensus ensemble learning through majority voting was used to determine the subset of AI methods with robust, reproducible performance with good generalization properties. The classifier aiming to predict the SD/(LD or LR) had a balanced accuracy of 88% (vs 81% for human readers consensus), a weighted precision of 94% (vs 87%), a weighted sensitivity of 94% (vs 88%) and specificity of 81% (vs 75%) and outperformed consensus of human readers (Tables 6, 7). The AI full prognosis solution SD/ LD/ LR had a balanced accuracy of 71%, a weighted precision of 77%, a weighted sensitivity of 74% and specificity of 82% to provide full prognosis (Figure 4). Ablation studies (Table 8) have been performed demonstrating the strong interest/added value of CT-imaging derived features and their complementarity with the biological/clinical and associated comorbidities. Short-term negative outcomes (Figure 4) referred to older patients with higher levels of lymphocytes and cardiac problems. Differentiation between long-term positive and negative outcome was observable according to the level of volume/heterogeneity of disease, the presence of cardiomegaly and cardiac calcifications and the condition of the non-infected lungs.

**Table 7.**
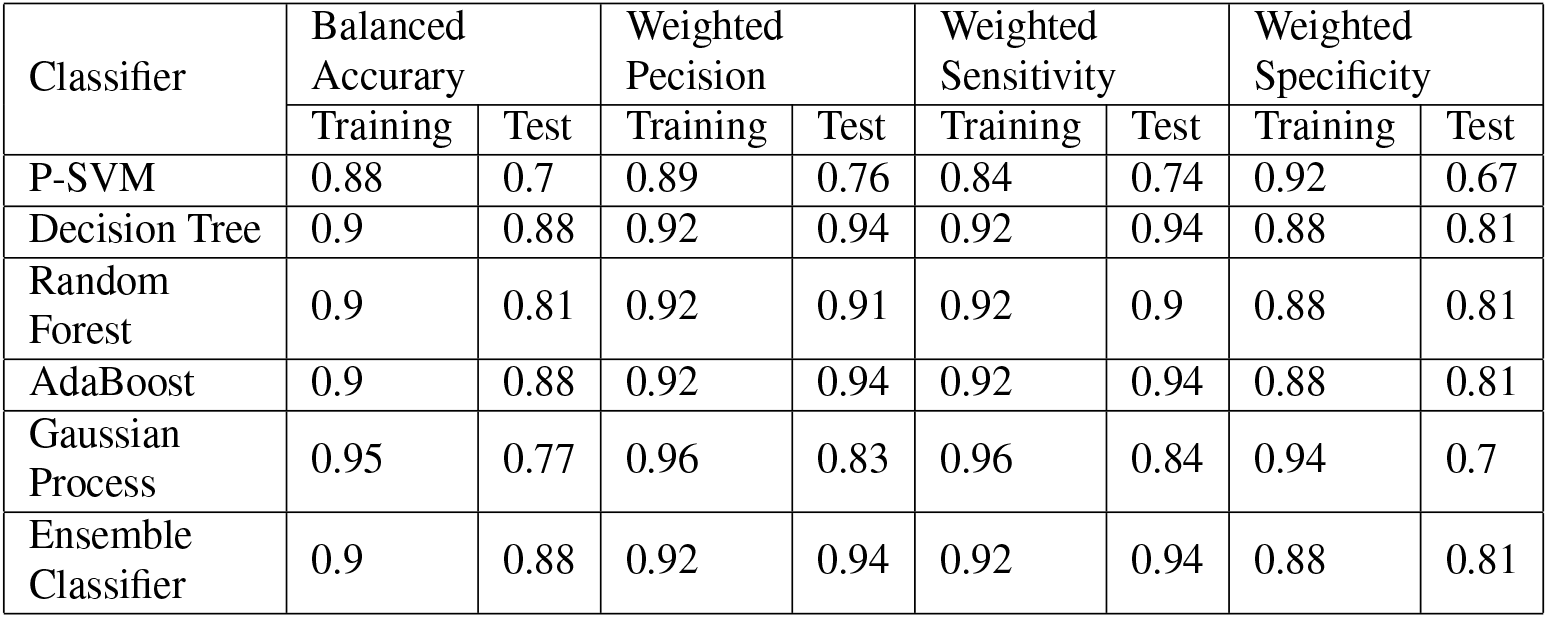
Performances of each of the top-5 individual classifiers and of the ensemble classifier to differentiate between Intubated (SI) and Deceased (SD) patients in the short-term outcome. *Note*.*- P-SVM = Support Vector Machine with a polynomial kernel*

**Table 8.** An ablation study of the different selected features. A leave-one-out method has been applied by removing one feature sequentially to test the features importance and the performance robustness. *Note*.*- a*) *D0: disease extent, b*) *D1: disease variables that are shape/geometry related, c*) *D2: disease variables that are tissue/texture, d*) *O1: heart/lungs variables that are shape/geometry related, e*) *O2: heart/lungs variables that are tissue/texture, f*) *B1: age, gender, biological/obesity/diabetes/fat/high blood pressure. LD = long-term-deceased; LR = long-term deceased; NS = non severe; S = severe; SI = short-term intubation; SD = short-term deceased*

| Study Case | Task | Balanced Accuracy |  | Weighted Precision |  | Weighted Sensitivity |  | Weighted Specificity |  |
| --- | --- | --- | --- | --- | --- | --- | --- | --- | --- |
|  |  | Training | Test | Training | Test | Training | Test | Training | Test |
| All Features | NS/S | 0.73 | 0.70 | 0.82 | 0.81 | 0.67 | 0.64 | 0.80 | 0.77 |
|  | SI/SD | 0.90 | 0.88 | 0.92 | 0.94 | 0.92 | 0.94 | 0.88 | 0.81 |
|  | LD/LR | 0.82 | 0.69 | 0.84 | 0.76 | 0.8 | 0.74 | 0.83 | 0.65 |
|  | SD/LD/LR | 0.77 | 0.71 | 0.8 | 0.77 | 0.78 | 0.74 | 0.9 | 0.82 |
| Without D0 | NS/S | 0.73 | 0.7 | 0.82 | 0.8 | 0.68 | 0.65 | 0.79 | 0.74 |
|  | SI/SD | 0.89 | 0.88 | 0.92 | 0.94 | 0.92 | 0.94 | 0.88 | 0.81 |
|  | LD/LR | 0.56 | 0.5 | 0.74 | 0.54 | 0.74 | 0.74 | 0.39 | 0.26 |
|  | SD/LD/LR | 0.65 | 0.58 | 0.73 | 0.64 | 0.76 | 0.74 | 0.79 | 0.72 |
| Without D1 | NS/S | 0.74 | 0.69 | 0.82 | 0.8 | 0.67 | 0.64 | 0.8 | 0.74 |
|  | SI/SD | 0.89 | 0.88 | 0.91 | 0.93 | 0.91 | 0.93 | 0.88 | 0.81 |
|  | LD/LR | 0.56 | 0.5 | 0.74 | 0.54 | 0.74 | 0.74 | 0.39 | 0.26 |
|  | SD/LD/LR | 0.65 | 0.58 | 0.73 | 0.64 | 0.76 | 0.74 | 0.79 | 0.72 |
| Without D2 | NS/S | 0.73 | 0.69 | 0.82 | 0.8 | 0.67 | 0.64 | 0.8 | 0.74 |
|  | SI/SD | 0.89 | 0.88 | 0.91 | 0.93 | 0.91 | 0.93 | 0.88 | 0.81 |
|  | LD/LR | 0.58 | 0.5 | 0.74 | 0.54 | 0.76 | 0.74 | 0.48 | 0.26 |
|  | SD/LD/LR | 0.67 | 0.58 | 0.73 | 0.64 | 0.76 | 0.74 | 0.82 | 0.72 |
| Without O1 | NS/S | 0.73 | 0.7 | 0.82 | 0.79 | 0.72 | 0.73 | 0.75 | 0.67 |
|  | SI/SD | 0.89 | 0.88 | 0.91 | 0.93 | 0.91 | 0.93 | 0.88 | 0.81 |
|  | LD/LR | 0.58 | 0.5 | 0.73 | 0.54 | 0.74 | 0.74 | 0.42 | 0.26 |
|  | SD/LD/LR | 0.66 | 0.58 | 0.72 | 0.64 | 0.76 | 0.74 | 0.81 | 0.72 |
| Without O2 | NS/S | 0.75 | 0.69 | 0.83 | 0.8 | 0.67 | 0.62 | 0.82 | 0.76 |
|  | SI/SD | 0.89 | 0.88 | 0.91 | 0.93 | 0.91 | 0.93 | 0.88 | 0.81 |
|  | LD/LR | 0.78 | 0.59 | 0.82 | 0.68 | 0.83 | 0.68 | 0.72 | 0.5 |
|  | SD/LD/LR | 0.74 | 0.65 | 0.78 | 0.73 | 0.79 | 0.7 | 0.87 | 0.78 |
| Without B1 | NS/S | 0.73 | 0.71 | 0.82 | 0.81 | 0.67 | 0.66 | 0.79 | 0.77 |
|  | SI/SD | 0.67 | 0.58 | 0.74 | 0.65 | 0.74 | 0.67 | 0.6 | 0.48 |
|  | LD/LR | 0.74 | 0.53 | 0.79 | 0.64 | 0.79 | 0.68 | 0.7 | 0.37 |
|  | SD/LD/LR | 0.58 | 0.41 | 0.59 | 0.48 | 0.59 | 0.48 | 0.73 | 0.66 |
| Clinical Only | NS/S | 0.71 | 0.58 | 0.8 | 0.73 | 0.68 | 0.58 | 0.73 | 0.58 |
|  | SI/SD | 0.89 | 0.88 | 0.91 | 0.93 | 0.91 | 0.93 | 0.88 | 0.81 |
|  | LD/LR | 0.73 | 0.53 | 0.79 | 0.64 | 0.8 | 0.68 | 0.65 | 0.37 |
|  | SD/LD/LR | 0.72 | 0.6 | 0.77 | 0.7 | 0.78 | 0.7 | 0.85 | 0.74 |

**Figure 4.**
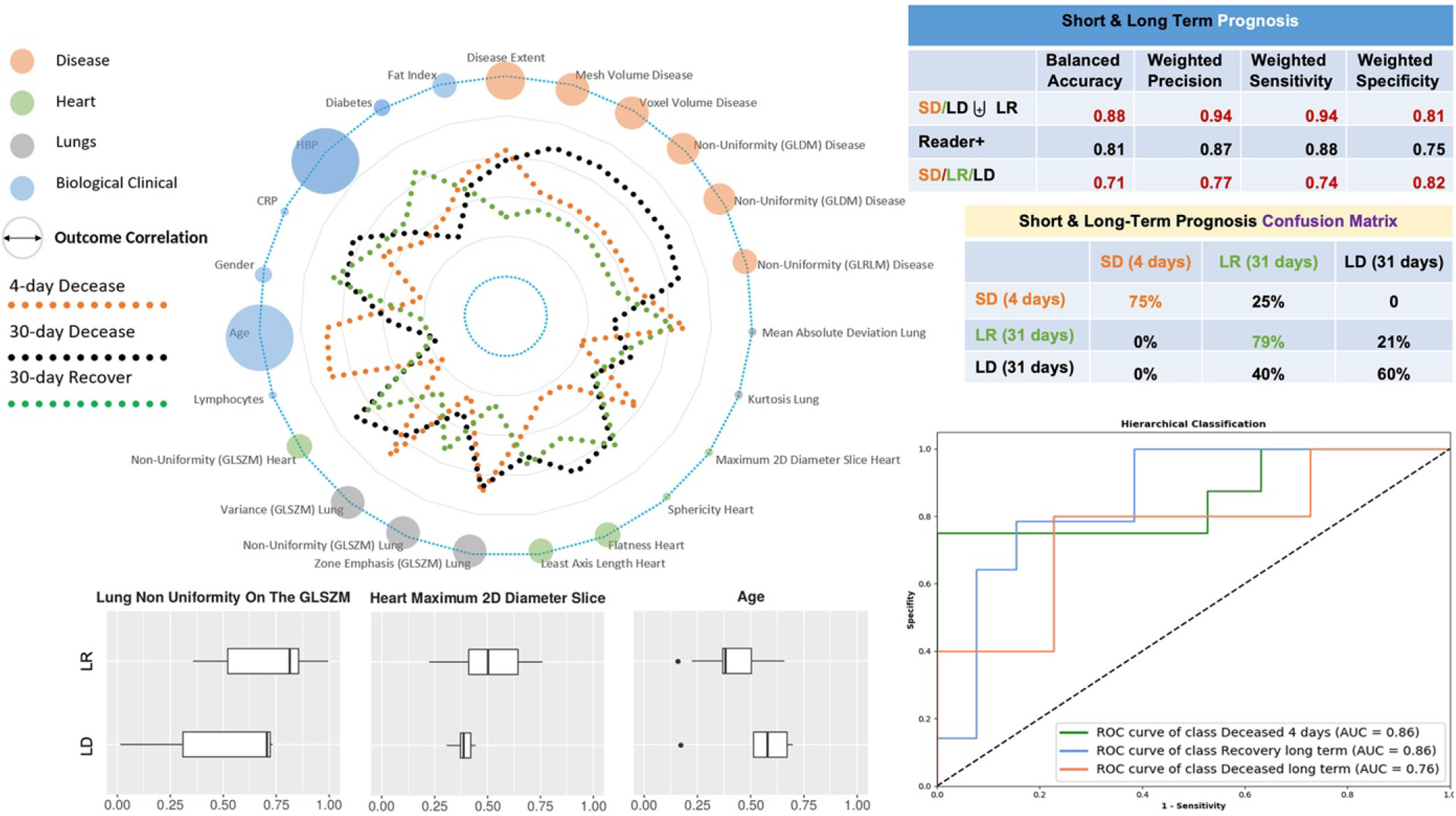
Short & Long Term Prognosis. Average profiles (spider chart) with respect to the short deceased (SD), long deceased (LD) and long recovered (LR) classes are shown along with their correlations with the outcome (diameter of the circle). Classification performance, confusion matrices and area under the curve with respect to the AI and - when feasible - the consensus of expert readers (reader+) are reported. Selective associations of features with final outcome are shown (box plots). ROC curves correspond to one-vs-all classification of the SD/LR/LD patients.

**Figure 5.**
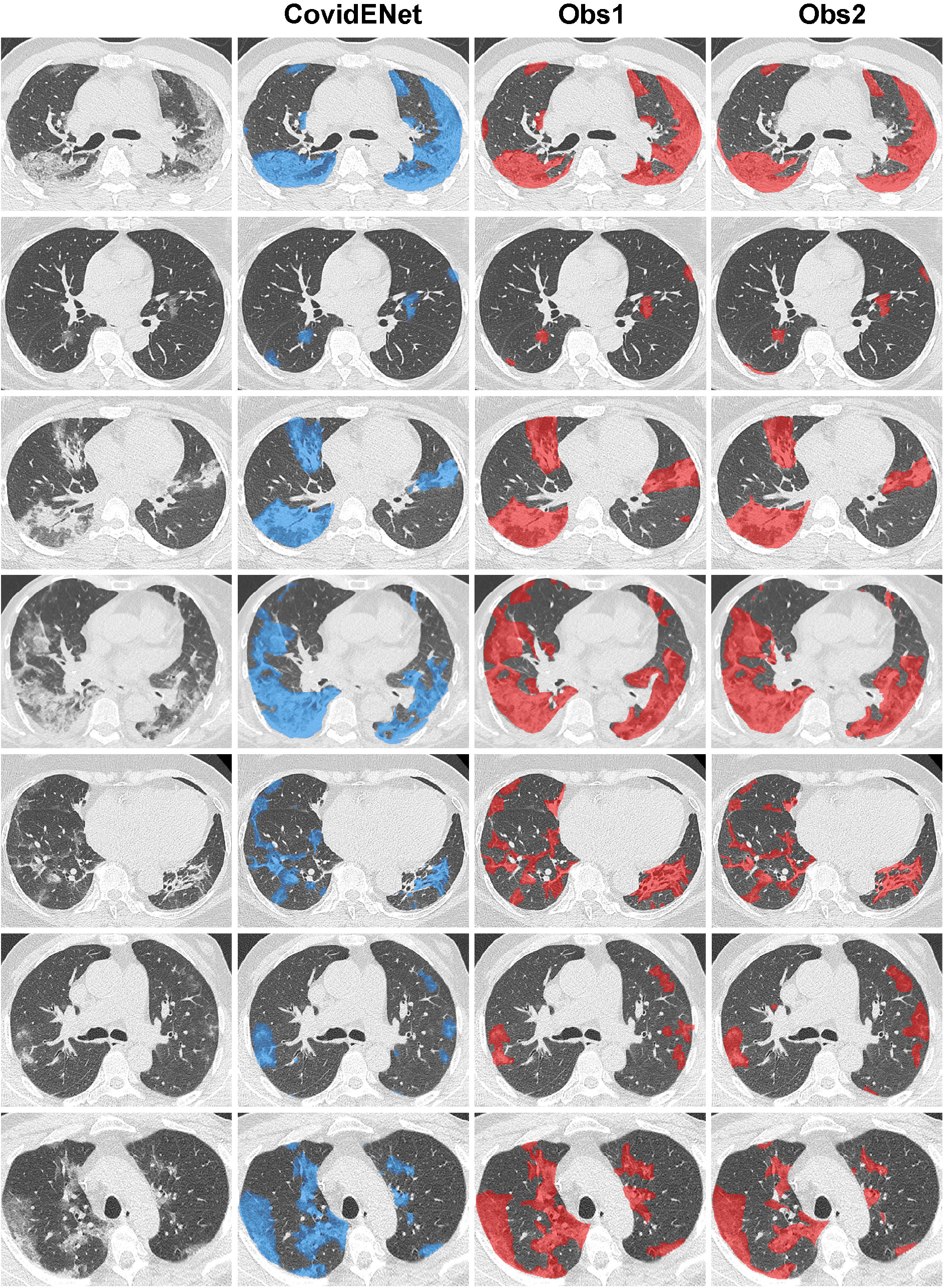
Some additional qualitative analysis for the comparison between automated and manual segmentations. Delineation of the diseased areas on chest CT in different slices of COVID-19 patients: From left to right: Input, AI-segmentation, expert I-segmentation, expert II-segmentation

### Part IV: Clinical Relevance & Impact

The pandemic requires implement rapid clinical triage in healthcare facilities to categorize patients according to urgency^24^, an objective our approach has fully addressed. Other studies have already reported on deep learning diagnosing COVID-19 pneumonia on chest radiograph^25^ or CT^16,26^, or quantifying it on CT^27,28^. Our study submits that AI should be part of triage, beyond the diagnostic value of biological, clinical and imaging data for COVID-19. To the best of our knowledge this study is the first to have developed a robust, holistic COVID19 multi-omics signature for disease staging and prognosis demonstrating an equivalent/superior-to-human-reader performance on a multi-centric data set. Our approach complied appropriate data collection and methodological testing requirements beyond the existing literature^26^. The proposed holistic signature harnessed imaging descriptors of disease, underlying lung, heart and fat as well as biological and clinical data. Among them, disease extent is known to be associated with severity^5,6^, disease textural heterogeneity reflects more the presence of heterogenous lesions than pure ground glass opacities observable in mild cases. Heart features encode cardiomegaly and cardiac calcifications. Lung features show patients with severe disease having greater dispersion and heterogeneity of lung densities, reflecting the presence of an underlying airway disease such as emphysema and the presence of sub-radiological disease. Among clinical variables, a higher CRP level, lymphopenia and a higher prevalence of hypertension and diabetes were associated with a poorer outcome, consistent with previous reports^4,9,10^. Interestingly, age was less predictive of disease severity than of poor outcome in severe patients. This is linked to the fewer therapeutic possibilities for these generally more fragile patients. Lastly, the average body mass index (BMI) in both non-severe and severe groups corresponded to overweight. Despite being correlated with BMI, the fat ratio measured on the CT scanner was only weakly associated with outcome. Several studies have reported obesity to be associated with severe outcomes^27,28^ and an editorial described the measurement of anthropometric characteristics as crucial to better estimate the risk of complications^19^. However a meta-analysis showed that whereas being associated with an increased risk of COVID-19 pneumonia, obesity was paradoxically associated with reduced pneumonia mortality^29^. Overall, the combination of clinical, biological and imaging features demonstrates their complementary value for staging and prognosis.

### Part V: Conclusions & Future Work

AI-enhanced imaging/clinical/biological/ information proved capable to identify patients with severe short/long-term outcomes, bolstering healthcare resources under the extreme pressure of the current COVID-19 pandemic. The information obtained from our AI staging and prognosis could be used as an additional element at admission to assist decision making. To conclude, our work contributes to addressing the pandemic in terms of (i) patient stratification with respect to the different therapeutic strategies, (ii) accelerated drug development through rapid, reproducible and quantified assessment of treatment response through the different mid/end-points of the trial, and (iii) continuous monitoring of patient response to treatment. In terms of future work, the continuous enrichment of the database with new examples is a necessary step on top of updating the outcome of patients included in the study. Furthermore, a fine-grained quantification of the disease depicting ground glass and consolidation could be of value both for staging and prognosis. The integration of D-dimer, that was available in a minority of patients is also an interesting future direction.

### Online Methods

#### Study Design and Participants

This retrospective multi-center study was approved by our Institutional Review Board (AAA-2020-08007) which waived the need for patients’ consent. Patients diagnosed with COVID-19 from March 4th to April 5th from eight large University Hospitals were eligible if they had positive reverse transcription polymerase chain reaction (PCR-RT) and signs of COVID-19 pneumonia on unenhanced chest CT. A total of 693 patients formed the full dataset (321, 360 CT slices). Only the CT examination performed at initial evaluation was included. Exclusion criteria were 1/ contrast medium injection and 2/ important motion artifacts. No patient was intubated at the time of the CT acquisition.

For the COVID-19 radiological pattern segmentation part, 50 patients from 3 centers (A: 20 patients; B: 15 patients, C: 15 patients) were included to compose a training and validation dataset, 130 patients from the remaining 3 centers (D: 50 patients; E: 50 patients, F: 30 patients) were included to compose the test dataset (Table 2). The patients from the training cohort were annotated slice-by-slice, while the patients from the testing cohort were partially annotated on the basis of 20 slices per exam covering in an equidistance manner the lung regions. The proportion between the CT manufacturers in the datasets was pre-determined in order to maximize the model generalizability while taking into account the data distribution.

For the staging (severe versus non-severe case) and prognosis (short and long-term) study, 513 additional patients from centers A (121 patients), B (157 patients), D (138 patients), G (77 patients) and H (20 patients) were included. Data of 536 patients from 5 centers (A, B, C, D and H) were used for training and those of 157 patients from 3 other centers (E, F and G) composed an independent test set (Table 3). Only one CT examination was included for each patient. Exclusion criteria were 1/ contrast medium injection and 2/ important motion artifacts. In addition to the CT examination - when available - patient sex, age, and body mass index (BMI), blood pressure and diabetes, lymphocyte count, CRP level and D-dimer level were also collected (Table 3).

For short-term outcome assessment, patients were divided into 2 groups: those who died or were intubated in the 4 days following the CT scan composed the severe short-term outcome subgroup, while the others composed the non-severe short-term outcome subgroup.

For long-term outcome, medical records were reviewed from May 7th to May 10th, 2020 to determine if patients died or had been intubated during the period of at least one month following the CT examination. The data associated with each patient (holistic profiling), as well as with the corresponding outcomes both in terms of severity assessment as well as in terms of final outcome and readers assessment will be made publicly available. Neither the CT scans, nor the associated experts’ or AI system annotations will be shared.

#### CT acquisitions

Chest CT exams were acquired on 4 different CT models from 3 manufacturers (Aquilion Prime from Canon Medical Systems, Otawara, Japan; Revolution HD from GE Healthcare, Milwaukee, WI; Somatom Edge and Somatom AS+ from Siemens Healthineer, Erlangen, Germany). The different acquisition and reconstruction parameters are summarized in Table 1. CT exams were mostly acquired at 120 (n=481*/*693; 69%) and 100 kVp (n=186*/*693; 27%). Images were reconstructed using iterative reconstruction with a 512×512 matrix and a slice thickness of 0.625 or 1 mm depending on the CT equipment. Only the lung images reconstructed with high frequency kernels were used for analysis. For each CT examination, dose length product (DLP) and volume Computed Tomography Dose Index (CTDIvol) were collected.

#### Data annotation

Fifteen radiologists (GC, TNHT, SD, EG, NH, SEH, FB, SN, CH, IS, HK, SB, AC, GF and MB) with 1 to 7 years of experience in chest imaging participated in the data annotation which was conducted over a 2-week period. For the training and validation set for the COVID-19 radiological pattern segmentation, the whole CT examinations were manually annotated slice by slice using the open source software ITKsnap ^1^. On each of the 23, 423 axial slices composing this dataset, all the COVID-19 related CT abnormalities (ground glass opacities, band consolidations, and reticulations) were segmented as a single class. Additionally, the whole lung was segmented to create another class (lung). To facilitate the collection of the ground truth for the lung anatomy, a preliminary lung segmentation was performed with Myrian XP-Lung software (version 1.19.1, Intrasense, Montpellier, France) and then manually corrected.

As far as test cohort for the segmentation is concerned, 20 CT slices equally spaced from the superior border of aortic arch to the lowest diaphragmatic dome were selected to compose a 2, 600 images dataset. Each of these images were systematically annotated by 2 out of the 15 participating radiologists who independently performed the annotation. Annotation consisted of manual delineation of the disease and manual segmentation of the lung without using any preliminary lung segmentation.

Additionally, 3 radiologists, an internationally recognized expert with 20+ years of experience in thoracic imaging (Reader^*A*^), a thoracic radiologist with 7+ years of experience (Reader^*B*^) and a resident with 6-month experience in thoracic imaging (Reader^*C*^) were asked to perform a triage (severe versus non-severe cases) and for the severe cases (short-term deceased versus short-term intubated) prognosis process to predict the short-term outcome.

#### Disease quantification

COVID-19 segmentation tool was built using an ensemble method combining a 2D & 3D neural network method. AtlasNet framework^20^, a 2D fully convolutional network (CovidE2D) with a SegNet-based architecture^22^ was coupled with a 3D fully convolutional network based on the 3D-UNet^21^ (CovidE3D). The AtlasNet framework combines a registration stage of the CT scans to a number of anatomical templates and consequently utilizes multiple deep learning-based classifiers trained for each template. At the end, the prediction of each model is back-projected to the original anatomy and a majority voting scheme is used to produce the final projection, combining the results of the different networks. A major advantage of the AtlasNet framework is that it incorporates a natural data augmentation by registering each CT scan to several templates. Moreover, the framework is agnostic to the segmentation model that will be utilized. For the registration of the CT scans to the templates, an elastic registration framework based on Markov Random Fields was used, providing the optimal displacements for each template^30^.

The architecture of the implemented CovidENet models was based on already established fully convolutional neural network designs from the literature^21,22^. Fully convolutional networks following an encoder decoder architecture both in 2D and 3D were developed and evaluated. For the CovidE2D models the CT scans were separated on the axial view. The network included 5 convolutional blocks, each one containing two Conv-BN-ReLU layer successions. Maxpooling layers were also distributed at the end of each convolutional block for the encoding part. Transposed convolutions were used on the decoding part to restore the spatial resolution of the slices together with the same successions of layers. For the CovidE3D, the model similarly consisted of five blocks with a down-sampling operation applied every two consequent Conv3D-BN-ReLU layers. Additionally, five decoding blocks were utilized for the decoding path, at each block a transpose convolution was performed in order to up-sample the input. Skip connections were also employed between the encoding and decoding paths. In order to train this model, cubic patches of size 64 × 64× 64 were randomly extracted within a close range of the ground truth annotation border in a random fashion. Corresponding cubic patches were also extracted from the ground truth annotation masks and the lung anatomy segmentation masks. To this end, we trained the model with the CT scan patch as input, the annotation patch as target and the lung anatomy annotation patch as a mask for calculating the loss function only within the lung region. In order to train all the models, each CT scan was normalized by cropping the Hounsfield units in the range [− 1024, 1000].

Regarding implementation details, 6 templates were used for the AtlasNet framework together with normalized cross correlation and mutual information as similarities metrics. The CovidE2D networks were trained using weighted cross entropy loss using weights depending on the appearance of each class and dice loss. Moreover, the CovidE3D network was trained using a dice loss.

The Dice loss (DL) and weighted cross entropy (WCE) are defined as follows:

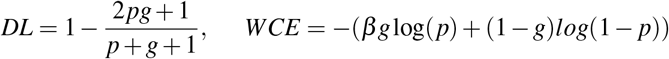

where *p* is the predicted from the network value and *g* the target/ ground truth value. *β* is the weight given for the less representative class. For network optimization, we used only the class for the diseased regions.

For the CovidE2D experiments we used classic stochastic gradient descent for the optimization with initial learning rate = 0.01, decrease of learning rate = 2.5 · 10^−3^ every 10 epochs, momentum =0.9 and weight decay =5 · 10^−4^. For CovidE3D experiments we used the AMSGrad and a learning rate of 0.001.

The training of a single network for both CovidE2D and CovidE3D network was completed in approximately 12 hours using a GeForce GTX 1080 GPU, while the prediction for a single CT scan was done in a few seconds. Training and validation curves for one template of CovidE2D and the CovidE3D networks are shown in Figure 6.

**Figure 6.**
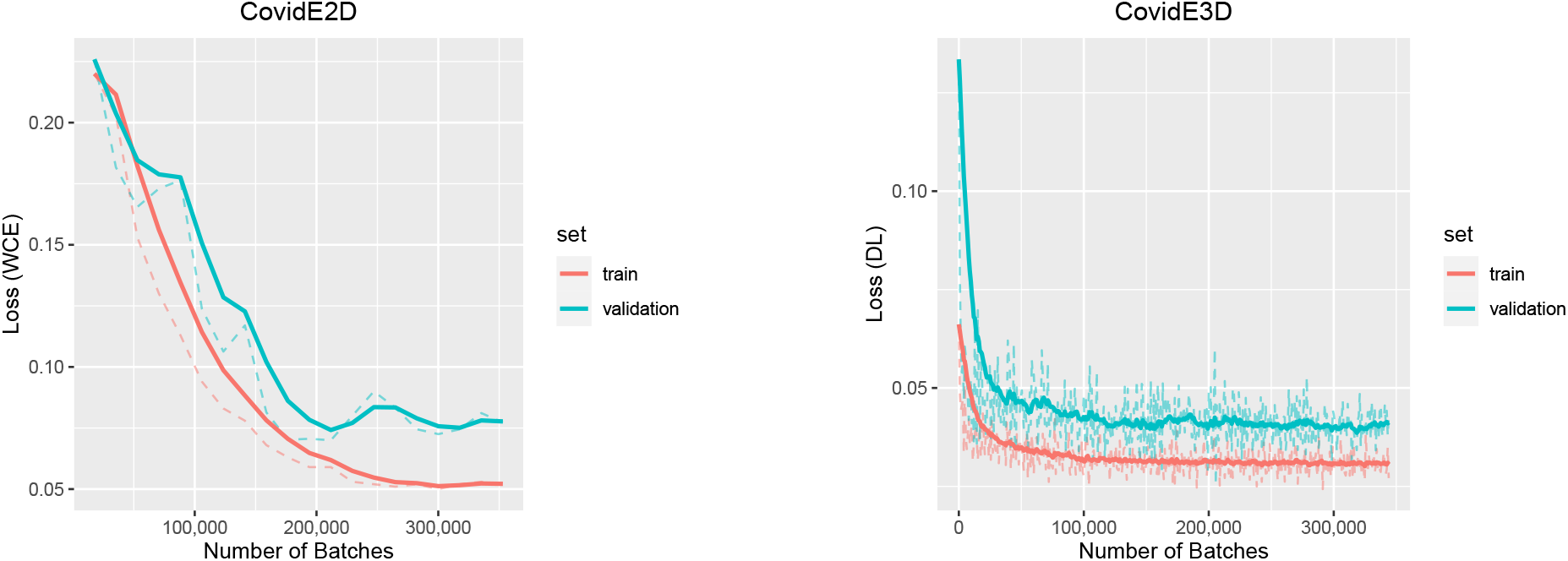
Training and validation curves for one template of AtlasNet and the 3D U-Net.

Both Dice similarity coefficient (DSC)^23^ and Haussdorff distances were higher with the 2D approach compared to the 3D approach (Figure 2). However, the combination of their probability scores led to a significant improvement. Thus, the ensemble of 2D and 3D architectures was selected for the final COVID-19 segmentation tool.

#### Lung/Breast/Heart & Body Contours Annotations

Lung, breast, heart and body contours segmentation masks of all patients were extracted by using ART-Plan software (TheraPanacea, Paris, France). ART-Plan is a CE-marked solution for automatic annotation of organs, harnessing a combination of anatomically preserving and deep learning concepts. This software has been trained using a combination of a transformation and an image loss. The transformation loss penalizes the normalized error between the prediction of the network and the affine registration parameters depicting the registration between the source volume and the whole body scanned. These parameters are determined automatically using a downhill simplex optimization approach. The second loss function of the network involved an image similarity function – the zero-normalized cross correlation loss – that seeks to create an optimal visual correspondence between the observed CT values of the source volume and the corresponding ones at the full body CT reference volume. This network was trained using as input a combination of 360, 000 pairs of CT scans of all anatomies and full body CT scans. These projections used to determine the organs being present on the test volume. Using the transformation between the test volume and the full body CT, we were able to determine a surrounding patch for each organ being present in the volume. These patches were used to train the deep learning model for each full body CT. The next step consisted of creating multiple annotations on the different reference spaces, and for that a 3D fully convolutional architecture was trained for every reference anatomy. This architecture takes as input the annotations for each organ once mapped to the reference anatomy and then seeks to determine for each anatomy a network that can optimally segment the organ of interest similar to the AtlasNet framework used for the disease segmentation. This information was applied for every organ of interest presented in the input CT Scan. In average, 6, 600 samples were used for training per organ after data augmentation. These networks were trained using a conventional dice loss. The final organ segmentation was achieved through a winner takes all approach over an ensemble networks approach. For each organ, and for each full body reference CT a specific network was built, and the segmentation masks generated for each network were mapped back to the original space. The consensus of the recommendations of the different subnetworks was used to determine the optimal label at the voxel level.

#### Holistic Multi-Omics Profiling & Staging

To this end, we investigate a variety of imaging characteristics extracted by the pathological regions as well as additional regions of interest that are associated with obesity, heart condition and healthy lung. These imaging characteristics are combined with powerful clinical and biological indicators that are reported from the literature to be associated with short and long-term outcome of Covid-19 patient progression.

##### Features extraction

A large variety of imaging features are extracted from the CT scans using the previously described segmentations of the disease, lung and heart. As a preprocessing step, all images were resampled by cubic interpolation to obtain isometric voxels with sizes of 1mm. Subsequently, disease, lung and heart masks were used to extract 107 radiomic features for each of them (left and right lung were considered separately both for the disease extent and entire lung). These features included first order statistics (maximum attenuation, skewness and 90th percentile etc), shape features (surface, maximum 2D diameter per slice, volume etc) and texture features (GLSZM, GLDM, GLRLM etc).

Two image indexes were also calculated, namely disease extent and fat ratio. The disease extent was calculated as the percentage of lung affected by the disease in respect to the entire lung volume. The disease components were extracted by calculating the number of individual connected components for the entire disease regions. Finally, as an indicator of obesity we calculated the fat ratio directly from the CT scans. The index was defined in an unsupervised manner. First the CT scans were smoothed using a Gaussian kernel with a standard deviation of 2 to remove noise and make the regions more homogeneous. Then, a threshold of the intensities in the range of [29, 130] was applied on the smoothed CTs indicating the fat regions. From these masks the regions of breast, lung and outside the body were excluded and the fat masks were calculated starting from the highest to the lowest part of the lungs. The fat index indicates the ratio of these masks on the corresponding body volume. To evaluate this morphometric measurement we assessed its correlation with BMI in the 362 patients for which BMI was available and found a strong correlation using Pearson correlation (r= 0.64; p<0.001; Figure 7).

**Figure 7.**
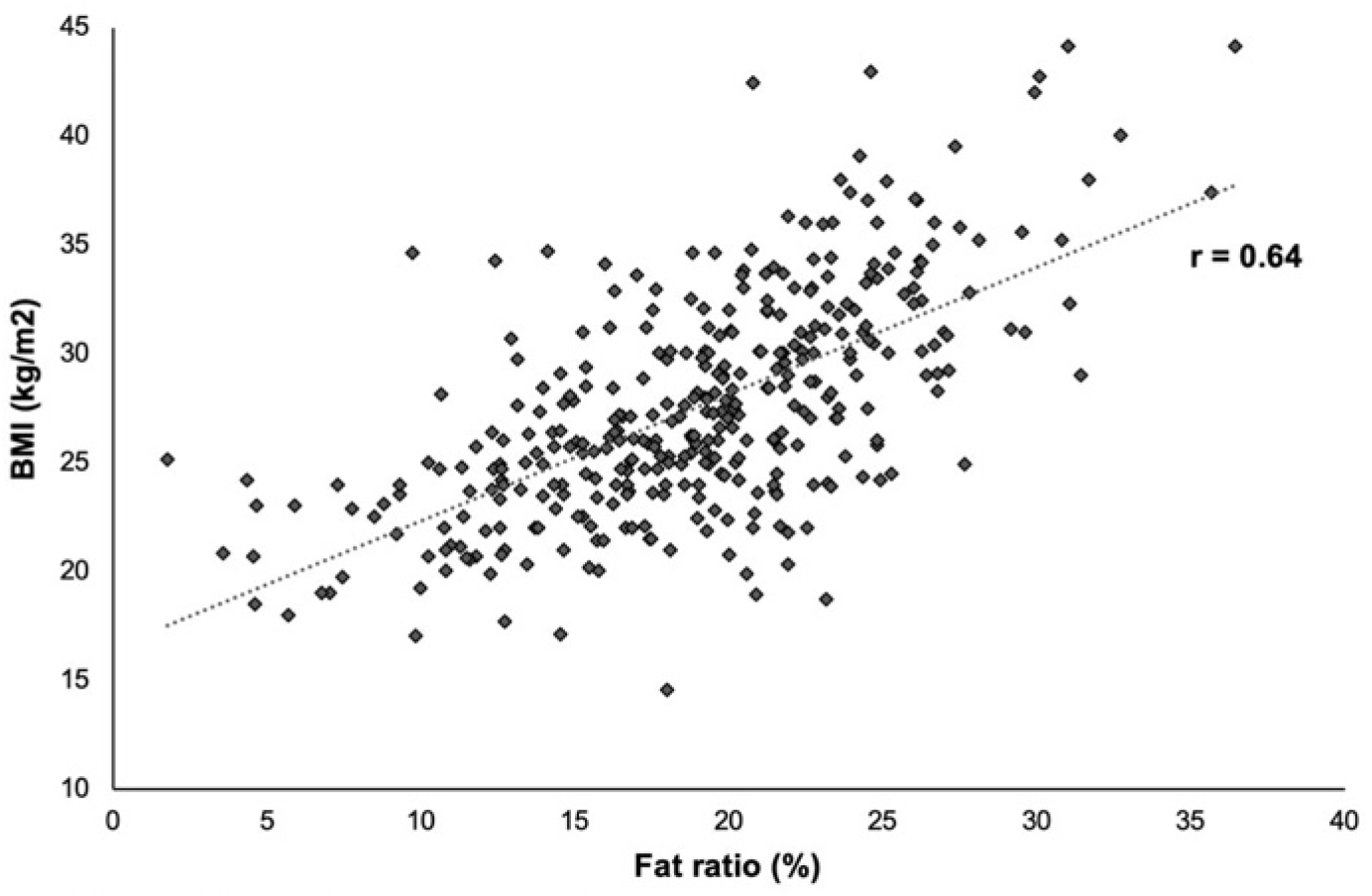
Correlation between body mass index (BMI) and fat ratio.

##### Holistic Biomarker Selection

Using all the calculated attributes (clinical, biological, imaging) we constructed a high dimensional space of size 543 - including clinical/biological variables -. A min-max normalization of the attributes was performed by calculating the minimum and maximum values for the training and validation cohorts. The same values were also applied on the test set.

To prevent overfitting and to also discover the most informative and robust attributes for the staging and prognosis of the patients we performed biomarker selection. Feature selection is very important for classification tasks and has been used widely in literature especially for radiomics^31^. First, the training data set was subdivided into training and validation on the principle of 80%-20% while respecting that the distribution of classes between the two subsets was identical to the observed one. To perform features selection, we have created 100 subdivisions on this basis and evaluated variety of classical machine learning - using the entire feature space - classifiers such as Decision Tree Classifier, Linear Support Vector Machine, XGBoosting, AdaBoost and Lasso. These classifiers were trained and validated to distinguish between Severe (S) and Non-Severe (NS) cases. In addition to these 5 classifiers-based feature selection approaches, we also considered statistics- based approaches based on Mutual Information, Chi-squared statistics and Univariate linear regression tests (Table 4). Each of these metrics was used to assess the correlation between the features and the outcome. Then, for each metric, the 5% features with the highest correlations were selected. Features were ranked according to their prevalence, the number of splits they were selected in, for each of the methods.

Our experiments indicated that different classifiers highlight different attributes as important. In order to take advantage of each feature selection properties, we adopted a consensus feature selection method. In particular, features having the best combined prevalence (sum of prevalences over the 8 selection techniques) were kept. For this feature selection task, Decision Tree Classifier was taken of maximum depth 3, Linear Support Vector Machine was taken with a linear kernel, a polynomial kernel function of degree 3 and a penalty parameter of 0.25, XGBoosting was used with a regression tree boosted over 30 stages, AdaBoost was used with a Decision Tree Classifier of maximum depth 2 boosted 3 times and Lasso method was used with 200 alphas along a regularization path of length 0.01 and limited to 1000 iterations.

##### Holistic COVID19 Multi-Omics Profiling

Using the descripted method, we have extracted 15 different features from the radiomics. These features belong to: features from imaging and in particular from the disease regions (5 features), lung regions (5 features) and heart features (5 features). To these radiomics features we added biological and clinical data (6 features: age, sex, high blood pressure (HBP), diabetes, lymphocyte count and CRP level) and image indexes (2 features: disease extent and fat ratio). At the end our biomarker consisted of 23 features in total. The correlation to outcome of these features is presented in the Table 9.

**Table 9.**
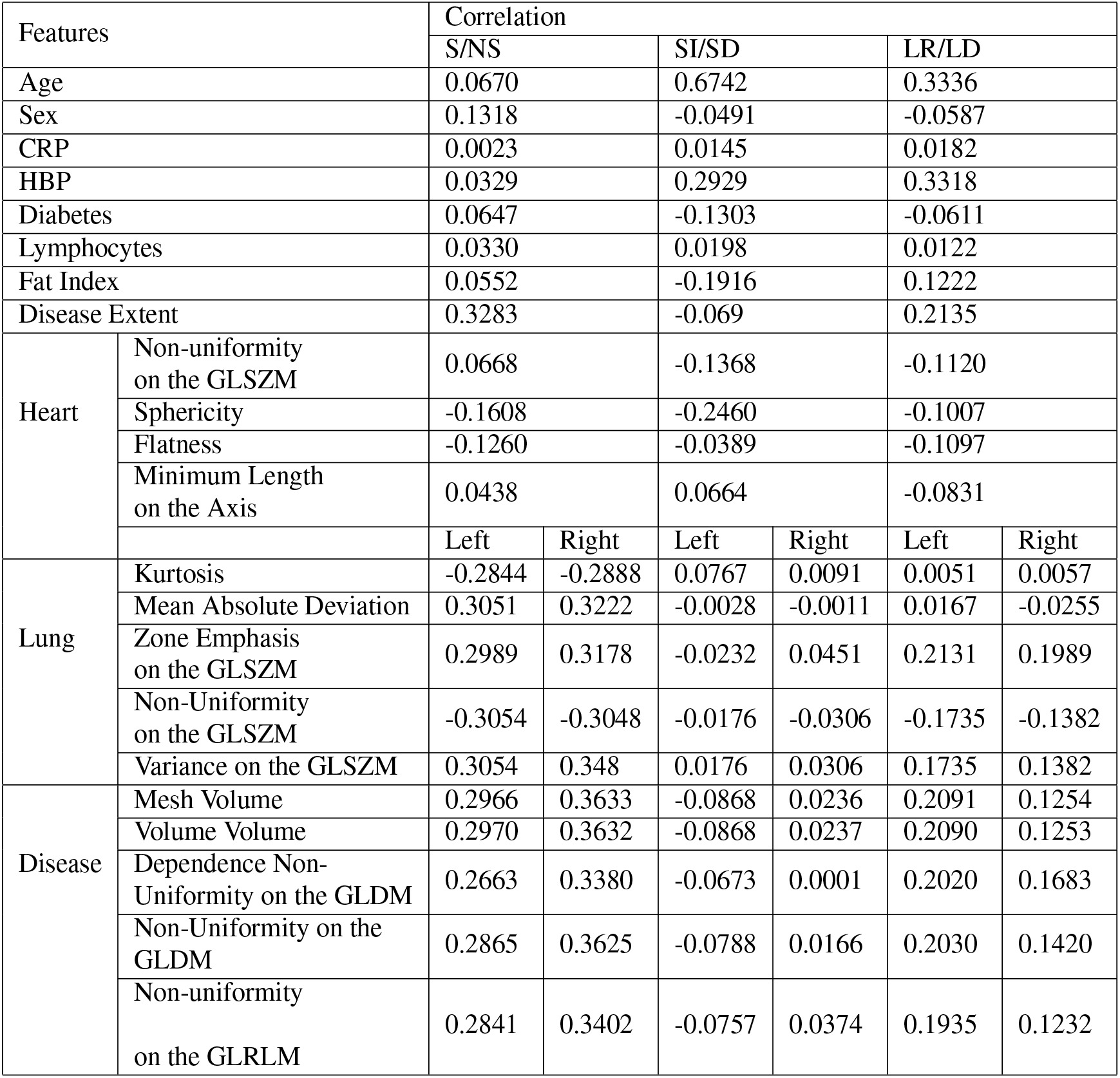
Correlation between outcome and the 23 selected features. *Note*.*— GLSZM =Gray Level Size Zone Matrix, GLRLM = Gray Level Run Length Matrix, GLDM = Gray Level Dependence Matrix, LD = long-term-deceased, LR = long-term deceased, NS = non severe, S = severe, SI = short-term intubation, SD = short-term deceased*

For the clinical and biological we kept all the collected features for our signature, except D-dimer level which was available in only 339*/*693 patients (Table 3). For other features missing data were imputed using the mean value on training set.

Regarding imaging features, we identified the following features as more important for the prognosis of the Covid-19 patients. These features include both first and second order statistics together with some shape features.

- Disease areas: Non-Uniformity of the Gray Level Dependence Matrix (GLDM), Dependence Non-Uniformity (GLDM), Mesh Volume, Voxel Volume, Non-Uniformity of the Gray level Run Length Matrix (GLRLM).
- Lung areas: Kurtosis, Mean Absolute Deviation, Zone Emphasis (GLSZM), Non-Uniformity (GLSZM), Variance (GLSZM).
- Heart areas: Maximum 2D diameter Slice, Non-Uniformity (GLSZM), Sphericity, Flatness, Minimum Length on the Axis.

The selected disease areas features capture both disease extent and disease textural heterogeneity. Disease textural heterogeneity is associated with lesions the presence of imaging pattern more complex than pure ground glass opacities usually found in mild disease. The selected lung areas features capture the dispersion and heterogeneity of lung densities, both of which may reflect the presence of an underlying airway disease such as emphysema but also the presence of sub-radiological disease. Lastly, the selected disease areas encode cardiomegaly and the presence of cardiac calcifications.

##### Staging Mechanism

The staging/prognosis component was addressed using an ensemble learning approach. Similarly to the biomarker extraction, the training data set was subdivided into training and validation sets on the principle of 80%-20%. This subdivision was performed such that the distribution of classes between the two subsets was identical to the observed one. We have created 10 subdivisions on this basis and evaluated the average performance of the following supervised classification methods: Nearest Neighbor, {Linear, Sigmoid, Radial Basis Function (RBF), Polynomial Kernel} Support Vector Machines (SVM), Gaussian Process, Decision Trees, Random Forests, AdaBoost, XGBoosting, Gaussian Naive Bayes, Bernoulli Naive Bayes, Multi-Layer Perceptron & Quadratic Discriminant Analysis. These classifiers have been trained using the identified holistic signature. For each binary classification task design a consensus model was designed selecting the top 5 classifiers with acceptable performance,> 60% in terms of balanced accuracy, as well as coherent performance between training and validation, performance decrease < 20% for the balanced accuracy between training and validation, were trained and combined together through a weighted winner takes all approach to determine the optimal outcome (Table 10, 11, 12). The weights granted to each selected classifier determined according to the rank of this classifier on validation regarding balanced accuracy weighted with a higher importance the best performing algorithms. Then, the selected classifiers were retrained using the entire training set, and their performance was reported in the external test cohort. Moreover, in order to assess the impact of each feature category to the implemented models we performed an ablation study by successively removing one category of features from the 6 categories defined for each classification task, results are presented in Table 8. The feature categories were identified as follow: a) D0: disease extent, b) D1: disease variables that are shape/geometry related, c) D2: disease variables that are tissue/texture, d) O1: heart/lungs variables that are shape/geometry related, e) O2: heart/lungs variables that are tissue/texture, B1: age, gender, biological/obesity/diabetes/fat/high blood pressure.

**Table 10.** Performances of the 15 evaluated classifiers for the Severe (S) versus Non-Severe (NS) task on cross-validation. *Note*.*-Asterixis indicates the top 5 classifiers reporting that were finally selected, L-SVM = Support Vector Machine with a linear kernel; P-SVM = Support Vector Machine with a polynomial kernel, S-SVM = Support Vector Machine with a sigmoid kernel, RBF-SVM = Support Vector Machine with a Radial Basis Function, QDA = Quadratic Discriminant Analysis*

| Classifier | Balanced Accuracy |  | Weighted Precision |  | Weighted Sensitivity |  | Weighted Specificity |  |
| --- | --- | --- | --- | --- | --- | --- | --- | --- |
|  | Training | Validation | Training | Validation | Training | Validation | Training | Validation |
| Nearest Neighbors | 0.73<br>±0.02 | 0.56<br>±0.03 | 0.86<br>±0.01 | 0.74<br>±0.03 | 0.87<br>±0.01 | 0.77<br>±0.01 | 0.59<br>±0.03 | 0.36<br>±0.05 |
| **L-SVM\*** | 0.71<br>±0.02 | 0.66<br>±0.05 | 0.8<br>±0.01 | 0.77<br>±0.03 | 0.72<br>±0.01 | 0.67<br>±0.06 | 0.71<br>±0.03 | 0.64<br>±0.05 |
| P-SVM | 0.79<br>±0.01 | 0.66<br>±0.03 | 0.85<br>±0.01 | 0.77<br>±0.02 | 0.8<br>±0.02 | 0.7<br>±0.04 | 0.78<br>±0.02 | 0.61<br>±0.04 |
| S-SVM | 0.57<br>±0.02 | 0.59<br>±0.03 | 0.72<br>±0.01 | 0.73<br>±0.02 | 0.61<br>±0.04 | 0.61<br>±0.02 | 0.53<br>±0.03 | 0.57<br>±0.06 |
| **RBF-SVM\*** | 0.75<br>±0.01 | 0.67<br>±0.03 | 0.83<br>±0.0 | 0.78<br>±0.02 | 0.7<br>±0.02 | 0.62<br>±0.05 | 0.8<br>±0.02 | 0.71<br>±0.04 |
| Gaussian Process | 0.62<br>±0.02 | 0.59<br>±0.04 | 0.82<br>±0.02 | 0.79<br>±0.04 | 0.83<br>±0.01 | 0.81<br>±0.02 | 0.41<br>±0.02 | 0.36<br>±0.06 |
| **Decision Tree\*** | 0.71<br>±0.01 | 0.7<br>±0.03 | 0.82<br>±0.01 | 0.82<br>±0.02 | 0.6<br>±0.03 | 0.6<br>±0.04 | 0.82<br>±0.02 | 0.81<br>±0.04 |
| **Random Forest\*** | 0.74<br>±0.01 | 0.68<br>±0.03 | 0.82<br>±0.01 | 0.79<br>±0.02 | 0.7<br>±0.02 | 0.66<br>±0.02 | 0.78<br>±0.02 | 0.71<br>±0.06 |
| Multi-Layer Perceptron | 0.98<br>±0.02 | 0.59<br>±0.03 | 0.99<br>±0.01 | 0.74<br>±0.02 | 0.99<br>±0.01 | 0.73<br>±0.04 | 0.97<br>±0.03 | 0.45<br>±0.03 |
| **AdaBoost\*** | 0.73<br>±0.01 | 0.69<br>±0.04 | 0.82<br>±0.01 | 0.8<br>±0.02 | 0.69<br>±0.07 | 0.66<br>±0.07 | 0.77<br>±0.06 | 0.73<br>±0.09 |
| **Gaussian Naive Bayes\*** | 0.63<br>±0.01 | 0.64<br>±0.05 | 0.78<br>±0.01 | 0.78<br>±0.03 | 0.81<br>±0.01 | 0.8<br>±0.03 | 0.45<br>±0.02 | 0.47<br>±0.07 |
| Bernoulli Naive Bayes | 0.52<br>±0.01 | 0.51<br>±0.01 | 0.85<br>±0.0 | 0.69<br>±0.05 | 0.81<br>±0.0 | 0.79<br>±0.01 | 0.24<br>±0.01 | 0.23<br>±0.02 |
| QDA | 0.78<br>±0.04 | 0.62<br>±0.04 | 0.86<br>±0.02 | 0.77<br>±0.03 | 0.85<br>±0.02 | 0.77<br>±0.03 | 0.71<br>±0.06 | 0.48<br>±0.08 |
| XGBoost | 0.7<br>±0.02 | 0.58<br>±0.02 | 0.9<br>±0.01 | 0.8<br>±0.02 | 0.88<br>±0.01 | 0.82<br>±0.01 | 0.52<br>±0.02 | 0.34<br>±0.04 |
| **Ensemble Classifier** | 0.75<br>±0.01 | 0.7<br>±0.03 | 0.83<br>±0.01 | 0.8<br>±0.02 | 0.69<br>±0.02 | 0.65<br>±0.03 | 0.8<br>±0.01 | 0.75<br>±0.05 |

**Table 11.** Performances of the 15 evaluated classifiers for the Intubated (SI) versus Deceased (SD) in the short-term task on cross-validation. *Note*.*-Asterixis indicates the top 5 classifiers reporting that were finally selected, L-SVM = Support Vector Machine with a linear kernel; P-SVM = Support Vector Machine with a polynomial kernel, S-SVM = Support Vector Machine with a sigmoid kernel, RBF-SVM = Support Vector Machine with a Radial Basis Function, QDA = Quadratic Discriminant Analysis*

| Classifier | Balanced Accuracy |  | Weighted Precision |  | Weighted Sensitivity |  | Weighted Specificity |  |
| --- | --- | --- | --- | --- | --- | --- | --- | --- |
|  | Training | Validation | Training | Validation | Training | Validation | Training | Validation |
| Nearest Neighbors | 0.81<br>±0.03 | 0.71<br>±0.07 | 0.88<br>±0.02 | 0.79<br>±0.05 | 0.88<br>±0.01 | 0.79<br>±0.04 | 0.75<br>±0.05 | 0.62<br>±0.11 |
| L-SVM | 0.8<br>±0.03 | 0.77<br>±0.06 | 0.83<br>±0.02 | 0.82<br>±0.05 | 0.76<br>±0.03 | 0.73<br>±0.05 | 0.83<br>±0.03 | 0.81<br>±0.08 |
| **P-SVM\*** | 0.86<br>±0.03 | 0.78<br>±0.05 | 0.88<br>±0.02 | 0.82<br>±0.03 | 0.82<br>±0.03 | 0.76<br>±0.05 | 0.89<br>±0.04 | 0.8<br>±0.06 |
| S-SVM | 0.63<br>±0.04 | 0.62<br>±0.09 | 0.73<br>±0.02 | 0.72<br>±0.07 | 0.74<br>±0.02 | 0.73<br>±0.05 | 0.51<br>±0.07 | 0.52<br>±0.14 |
| RBF-SVM | 0.7<br>±0.06 | 0.64<br>±0.07 | 0.84<br>±0.01 | 0.77<br>±0.08 | 0.83<br>±0.02 | 0.78<br>±0.05 | 0.57<br>±0.1 | 0.5<br>±0.09 |
| Gaussian Process | 0.92<br>±0.05 | 0.84<br>±0.05 | 0.95<br>±0.03 | 0.88<br>±0.04 | 0.95<br>±0.03 | 0.87<br>±0.05 | 0.9<br>±0.06 | 0.81<br>±0.08 |
| **Decision Tree\*** | 0.89<br>±0.03 | 0.89<br>±0.08 | 0.91<br>±0.02 | 0.91<br>±0.06 | 0.91<br>±0.03 | 0.89<br>±0.08 | 0.88<br>±0.04 | 0.88<br>±0.09 |
| **Random Forest\*** | 0.9<br>±0.04 | 0.83<br>±0.11 | 0.93<br>±0.03 | 0.88<br>±0.08 | 0.92<br>±0.04 | 0.87<br>±0.07 | 0.88<br>±0.05 | 0.79<br>±0.15 |
| Multi-Layer Perceptron | 1.0<br>±0.01 | 0.79<br>±0.08 | 1.0<br>±0.0 | 0.84<br>±0.05 | 1.0<br>±0.0 | 0.83<br>±0.04 | 1.0<br>±0.01 | 0.75<br>±0.13 |
| **AdaBoost\*** | 0.89<br>±0.03 | 0.88<br>±0.08 | 0.91<br>±0.02 | 0.91<br>±0.06 | 0.91<br>±0.02 | 0.89<br>±0.08 | 0.88<br>±0.05 | 0.86<br>±0.09 |
| Gaussian Naive Bayes | 0.66<br>±0.05 | 0.62<br>±0.08 | 0.81<br>±0.03 | 0.77<br>±0.1 | 0.81<br>±0.02 | 0.78<br>±0.05 | 0.52<br>±0.09 | 0.47<br>±0.12 |
| Bernouilli Naive Bayes | 0.59<br>±0.02 | 0.53<br>±0.03 | 0.83<br>±0.01 | 0.66<br>±0.14 | 0.79<br>±0.01 | 0.76<br>±0.02 | 0.39<br>±0.03 | 0.3<br>±0.05 |
| QDA | 0.61<br>±0.06 | 0.5<br>±0.0 | 0.82<br>±0.03 | 0.55<br>±0.0 | 0.8<br>±0.03 | 0.74<br>±0.0 | 0.43<br>±0.09 | 0.26<br>±0.0 |
| **XGBoost\*** | 0.97<br>±0.02 | 0.78<br>±0.13 | 0.99<br>±0.01 | 0.84<br>±0.1 | 0.99<br>±0.01 | 0.83<br>±0.09 | 0.96<br>±0.03 | 0.74<br>±0.19 |
| **Ensemble Classifier** | 0.93<br>±0.03 | 0.87<br>±0.08 | 0.94<br>±0.02 | 0.9<br>±0.06 | 0.93<br>±0.02 | 0.89<br>±0.07 | 0.92<br>±0.04 | 0.85<br>±0.12 |

**Table 12.**
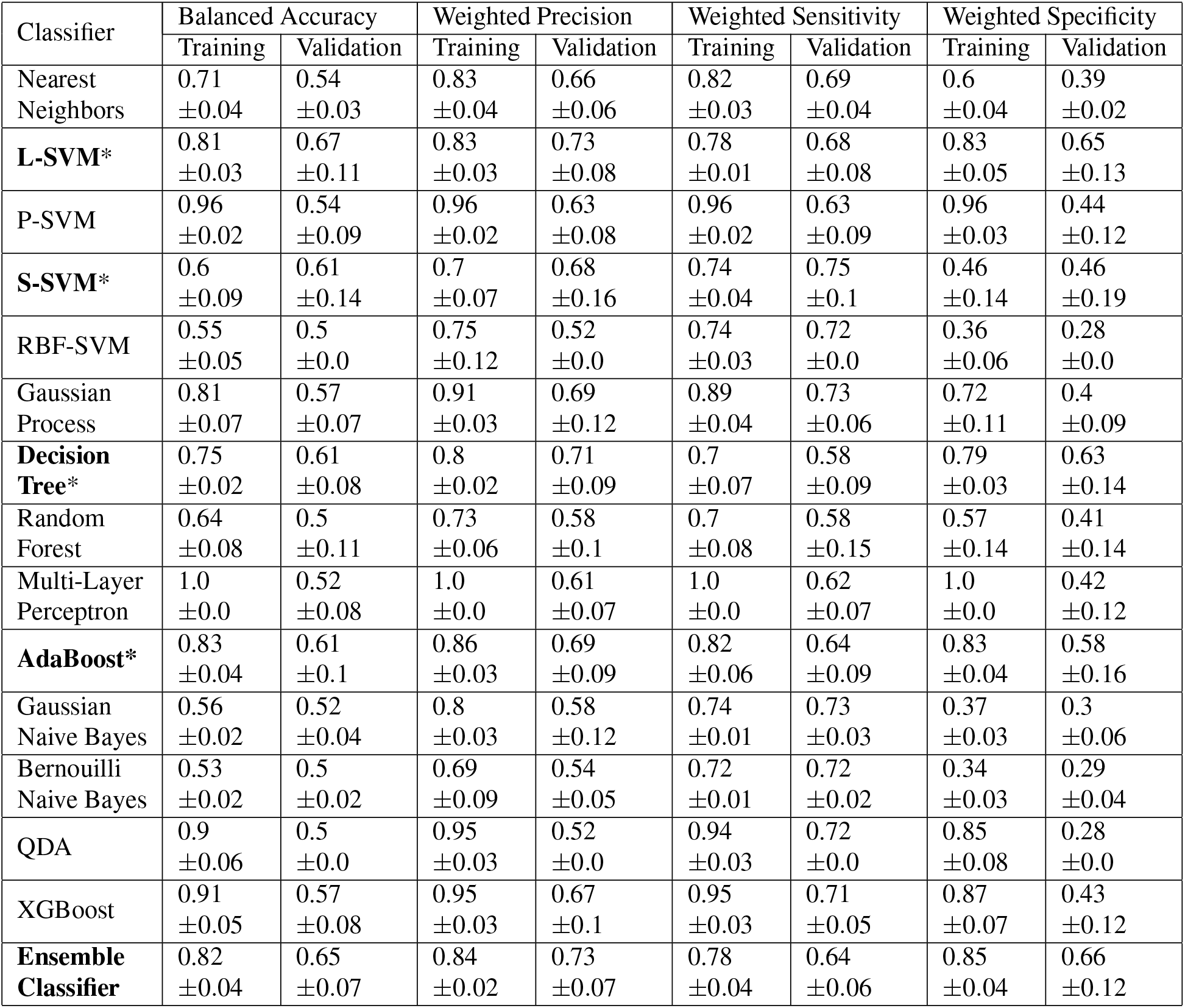
Performances of the 15 evaluated classifiers for the Deceased (LD) and Recovered (LR) in the long-term task on cross-validation. *Note*.*-Asterixis indicates the top 5 classifiers reporting that were finally selected, L-SVM = Support Vector Machine with a linear kernel; P-SVM = Support Vector Machine with a polynomial kernel, S-SVM = Support Vector Machine with a sigmoid kernel, RBF-SVM = Support Vector Machine with a Radial Basis Function, QDA = Quadratic Discriminant Analysis*

##### Prognosis Mechanism

To perform the Short-term deceased (SD)/ long-term Deceased (LD)/Long term recovered (LR) classification task, a SD/SI (SI: Intubated at 4 days) classifier and a LD/LR classifiers were applied in a hierarchical way, performing first the short-term staging and then the long-term prognosis for patients classified as in need of mechanical ventilation support. More specifically, a majority voting method was applied to classify patients into SD and SI cases (Table 11). Then, another hierarchical structure was applied on the cases predicted as SI only to classify them into the ones who didn’t recover within 31+ days of mechanical ventilation (LD) and the ones who recovered with 30 days on mechanical ventilation (LR). The ensemble of the classifiers was trained using the same technique (Table 12).

Concerning the implementation details, to overcome the unbalance dataset for the different classes, each class received a weight inversely proportional to its size. For the NS versus S majority voting classifier the top 5 classifiers consists in RBF SVM, Linear SVM, AdaBoost, Random forest, Decision Tree (Table 5). The SVM methods were granted a polynomial kernel function of degree 3, the Linear kernel one had a penalty parameter of 0.3 while the RBF SVM had a penalty parameter of 0.15. In addition, the RBF SVM was granted a kernel coefficient of 1. The Decision Tree classifier was limited to a depth of 2 to avoid overfitting. The Random Forest classifier was composed of 25 of such Decision Trees. AdaBoost classifier was based on a decision tree of maximal depth of 1 boosted 4 times. For the SI versus SD majority voting classifier the top 5 classifiers consists in polynomial SVM, Linear SVM, Decision Tree, Random Forest and AdaBoost (Table 7). The Linear and Polynomial SVM were granted a polynomial kernel function of degree 2 and a penalty parameter of 0.35. The Decision Tree classifier was limited to a depth of 1 and Random Forest was composed of 50 of such trees. AdaBoost classifier was based on a decision tree of maximal depth of 1 boosted 2 times. Finally, the LR versus LD (Table 13) majority voting classifier was only using the 4 classifiers with balanced accuracy > 0.6 namely Linear and Sigmoid SVM, Decision Tree, and AdaBoost Classifiers. The SVM methods were defined with a kernel function of degree 3 and a penalty parameter of 1. Decision Tree was defined to a depth of 1, AdaBoost being defined with such a Decision Tree boosted 3 times. In the extended Figures 8, 9, 10 we visualize the distributions of the different features along the ground truth labels of the hierarchical classifier for each subject. In particular, all the samples are grouped using their ground truth labels and a boxplot is generated for each group and each feature. The boxes had been generated by using the 25th, median and 75th percentile of the features. It is therefore clearly visible that some features such as the disease extent, the age, the shape of the disease and the uniformity seems to be very important on separating the different subjects.

**Table 13.** Performances of each of the individual classifiers and of the ensemble classifier to differentiate between Deceased (LD) and Recovered (LR) in the long-term outcome. *Note*.*- P-SVM = Support Vector Machine with a polynomial kernel; S-SVM = Support Vector Machine with a sigmoid kernel*

| Classifier | Balanced Accuracy |  | Weighted Precision |  | Weighted Sensitivity |  | Weighted Specificity |  |
| --- | --- | --- | --- | --- | --- | --- | --- | --- |
|  | Training | Test | Training | Test | Training | Test | Training | Test |
| L-SVM | 0.77 | 0.62 | 0.81 | 0.7 | 0.74 | 0.63 | 0.81 | 0.61 |
| S-SVM | 0.63 | 0.69 | 0.71 | 0.76 | 0.56 | 0.63 | 0.7 | 0.74 |
| AdaBoost | 0.82 | 0.69 | 0.84 | 0.76 | 0.8 | 0.74 | 0.83 | 0.65 |
| Decision Tree | 0.7 | 0.72 | 0.8 | 0.78 | 0.6 | 0.68 | 0.81 | 0.76 |
| Ensemble Classifier | 0.82 | 0.69 | 0.84 | 0.76 | 0.8 | 0.74 | 0.83 | 0.65 |

**Figure 8.**
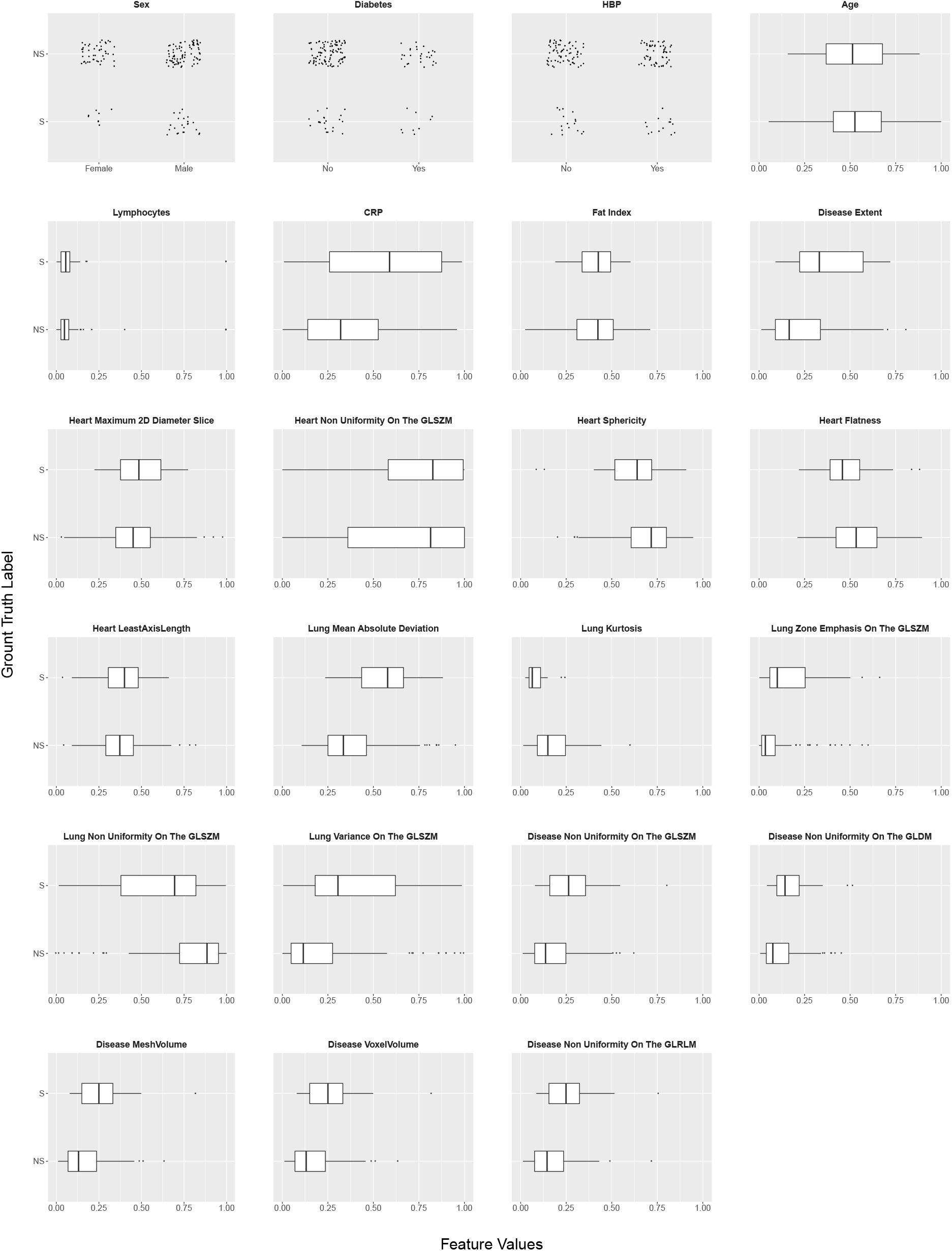
Boxplots of the selected features and their association with the annotation labels for the severe (S) versus non severe (NS) cases on the short-term outcome for the test set. The boxes had been generated by using the 25th, median and 75th percentile of the features.

**Figure 9.**
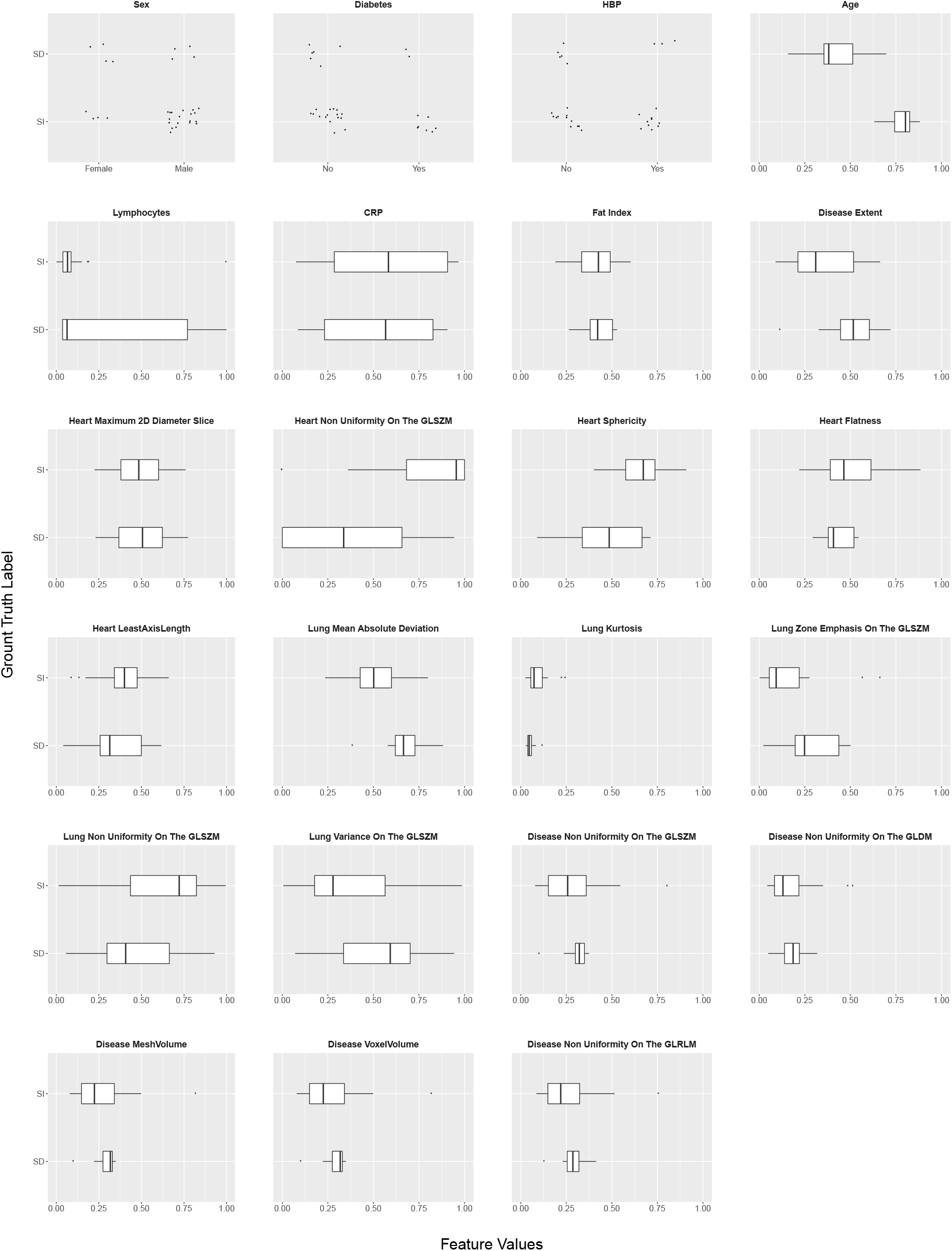
Boxplots of the selected features and their association with the annotation labels for the short intubated (SI) versus deceased (SD) on the short-term outcome for the test set. The boxes had been generated by using the 25th, median and 75th percentile of the features.

**Figure 10.**
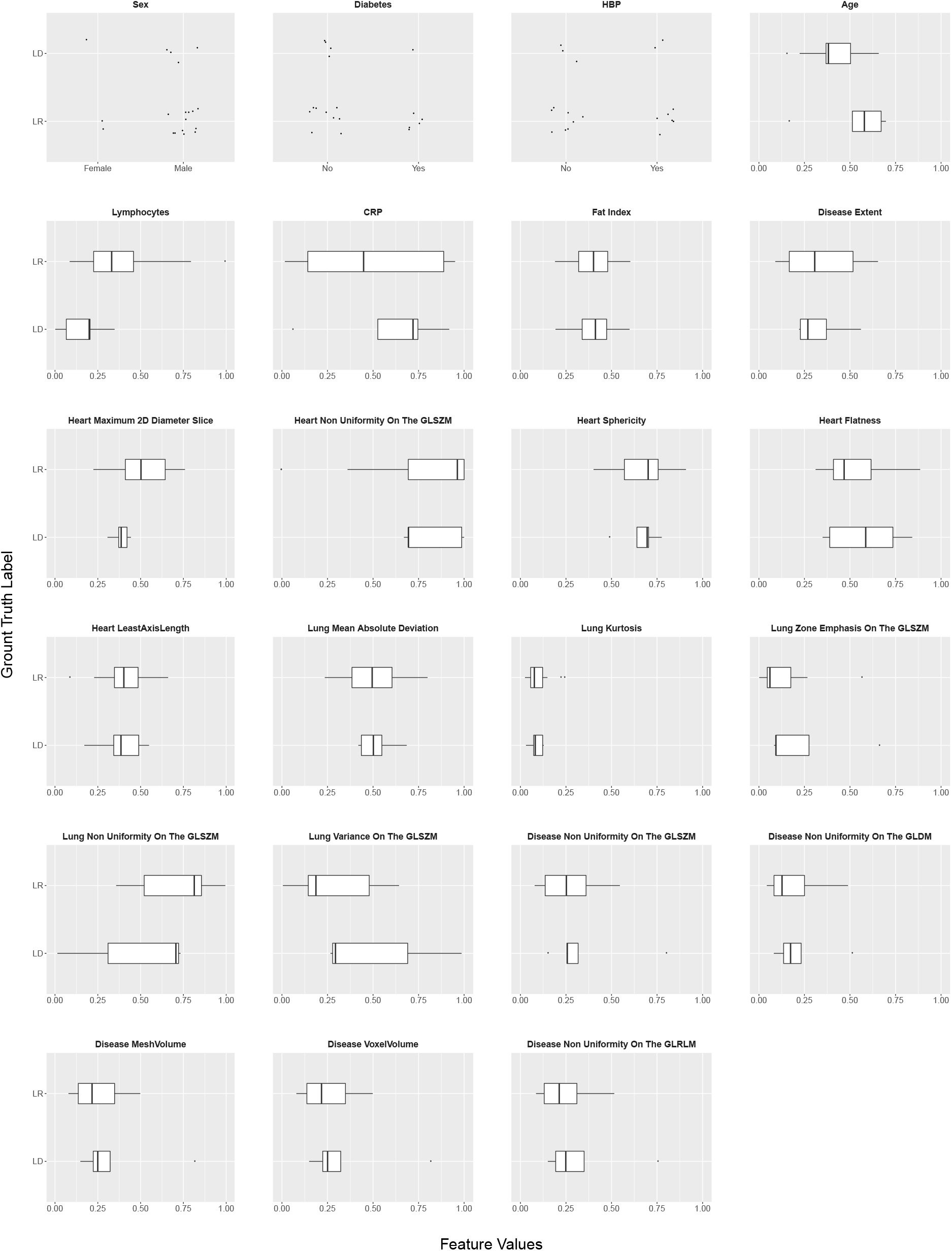
Boxplots of the selected features and their association with the ground truth labels for the recovered (LR) versus deceased (LD) on the long-term outcome for the test set. The boxes had been generated by using the 25th, median and 75th percentile of the features.

##### Statistical Analysis

The statistical analysis for the deep learning-based segmentation framework and the radiomics study was performed using Python 3.7, Scipy^32^, Scikit-learn^33^, TensorFlow^34^ and Pyradiomics^35^ libraries. The dice similarity score (DSC)^23^ was calculated to assess the similarity between the 2 manual segmentations of each CT exam of the test dataset and between manual and automated segmentations. The DSC between manual segmentations served as reference to evaluate the similarity between the automated and the two manual segmentations. Moreover, the Hausdorff distance was also calculated to evaluate the quality of the automated segmentations in a similar manner. Disease extent was calculated by dividing the volume of diseased lung by the lung volume and expressed in percentage of the total lung volume. Disease extent measurement between manual segmentations and between automated and manual segmentations were compared using paired Student’s t-tests.

For the stratification of the dataset into the different categories, classic machine learning metrics, namely balanced accuracy, weighted precision, and weighted specificity and sensitivity were used. The weighted metrics are expressed as follows. If we denote *N* the total number of samples, *N*_*l*_ the number of samples with class of label *l* and *S*_*l*_ the non-weighted score in one-vs-rest classification for class of label *l*, the corresponding weighted score WS is then:

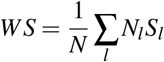

Moreover, the correlations between each feature and the outcome was computing using a Pearson correlation over the entire dataset.Patient characteristics between training/validation and test datasets were compared using chi-square and Student’s t-tests.

##### Author Contributions

GC, MV, MPR and NP designed the study protocol

GC, SD, EG, FB, SN, IS, HK, LF, HM, TG, JG, YN, AK, EM, PYB, STB, VB, AM, RYC and MPR performed data acquisition

GC, TNHT, SD, EG, NH, SEH, FB, SN, CH, IS, HK, SB, AC, GF and MB performed data annotation MV, EB, SC, ED, FP, AL and NP performed data analysis

GC, MV, MPR and NP drafted the manuscript. All the authors provided scientific input, revised and approved the manuscript.

## Data Availability

N/A

## Footnotes

1 http://www.itksnap.org

